# White matter alterations in focal to bilateral tonic-clonic seizures

**DOI:** 10.1101/2021.10.21.21265249

**Authors:** Christina Maher, Arkiev D’Souza, Rui Zeng, Michael Barnett, Omid Kavehei, Armin Nikpour, Chenyu Wang

## Abstract

We sought to examine the microstructural white matter differences in patients with focal to bilateral tonic-clonic seizures (FBTCS), compared to those with focal epilepsy without FBTCS, and control participants. Using a superior tract segmentation model, we obtained track-weighted tensor-metrics (TW-TM), implemented through an automated pipeline for image analysis and tract reconstruction. Analysis of covariance was used to compare group differences in the TW-TM for whole-tract and hemispheric tract measurements. We identified several white matter regions that displayed significantly altered white matter in patients with focal epilepsy compared to controls. Further, patients without FBTCS had significantly increased white matter disruption in the inferior fronto-occipital fascicle and the striato-occipital tract. In contrast, patients with FBTCS were more similar to healthy controls in most regions, except for distinct alterations in the inferior cerebellar region compared to the non-FBTCS group and controls. Our study revealed marked alterations in a range of subcortical tracts widely considered critical in the genesis of seizures in focal epilepsy. Our application of TW-TM in a new clinical dataset enabled the identification of specific tracts that may act as a predictive biomarker to distinguish patients who are likely to develop FBTCS.

## 1 INTRODUCTION

For drug-resistant patients with focal to bilateral tonic-clonic seizures (FBTCS), there is an increased risk of cardiac arrhythmias^1^, seizure-related injuries^2^ and sudden unexpected death in epilepsy (SUDEP)^3^. Unlike other epilepsies, such as temporal lobe epilepsy (TLE), where the role of structural regions is largely understood, the mechanisms underlying FBCTS remain elusive. Therefore, the importance of delineating the mechanisms involved in FBTCS is amplified as a vital objective to aid control of FBTCS and prevention of SUDEP.

Although the taxonomy of FBTCS implies whole-brain generalisation of seizures, FBTCS are primarily highly selective, producing more vigorous activity in specific brain regions^4,5,6^. Mounting evidence has endorsed the subcortical structures such as the thalamus and basal ganglia (BG) and their associated circuits as critical to the information relay involved in FBTCS. The thalamocortical relay fibres are topographically arranged to project to the cerebral cortex, from which sensory information is processed and relayed back to the original projection site in the thalamus. Acting as a “relay station”^7^, the thalamus has widespread connections across the entire cerebral cortex^8^ and moderates communication between various brain regions. Within the context of FBTCS, the thalamus has been proposed as a support system for seizure propagation via its role in the synchronisation of abnormal cortical-subcortical ictal discharge^9,10^.

On the other hand, the BG functions as a “braking system”, interacting with the thalamus and cortex through multiple parallel circuits, including the direct and indirect pathways^11^. The BG is increasingly hypothesised to play an anticonvulsive role in FBTCS^12^, yet specific mechanisms remain unclear. Increased BG activity was reported to be negatively associated with FBTCS in TLE^13,14^. In contrast, others illustrated that the BG only become involved when ictal activity disperses to additional cortical regions during secondary generalisation^15^.

The subcortical structures involved in FBTCS are also involved in focal seizures^16^. We included both patient groups (focal with FBTCS, termed “FBTCS-Y”, and focal only, termed “FBTCS-N”) in this study to address the distinct structural differences between the two groups. In addition to the thalamic and striatal regions, we included other white matter regions based on their well-documented role in focal seizures and functional connectivity^17,18^, to determine whether any observed group differences were unique to the FBTCS group, and to account for possible whole brain differences.

Tensor-based metrics derived from diffusion tensor imaging (DTI) have been used to identify microstructural white matter alterations in a range of epilepsies^19,20^. Meta-analysis showed patients with focal epilepsy had elevated regional mean diffusivity relative to controls^20^. However, FA findings are less consistent; some studies report reduced FA in patients with epilepsy compared to controls^18,21^, whilst others report no change across various regions^22,23^; thus, further clarification is required. Moreover, though tensor-based metrics have been established as a valuable technique to elucidate the mechanisms of focal epilepsy, they have seldom been applied to explore the FBTCS patient population.

This study measured tensor-based metrics to explore new biomarkers in patients with focal epilepsy and FBTCS. We hypothesised that compared to controls, (1) patients with focal epilepsy (“All patients” group) would have significantly altered white matter in a range of subcortical regions; (2) the patients with FBTCS would have significantly altered white matter in the thalamic and striatal regions compared to those without FBTCS.

## 2 RESULTS

### 2.1 Demographics

Twenty-five patients (10M, 15F, mean age 40 ±12.7 years, 17 with FBTCS-Y and 8 with FBTCS-N) and 19 controls (5M, 15F, mean age 37 ±11.12 years) were included in this study after passing the imaging quality control (QC) check. There were no significant differences in age or gender in both grouping conditions (“All patients” versus controls; FBTCS-Y, FBTCS-N versus controls). There was no significant association between lesion presence and seizure onset side; or lesion presence and FBTCS. Table 1 shows the characteristics of the patients and control participants.

**TABLE 1.**
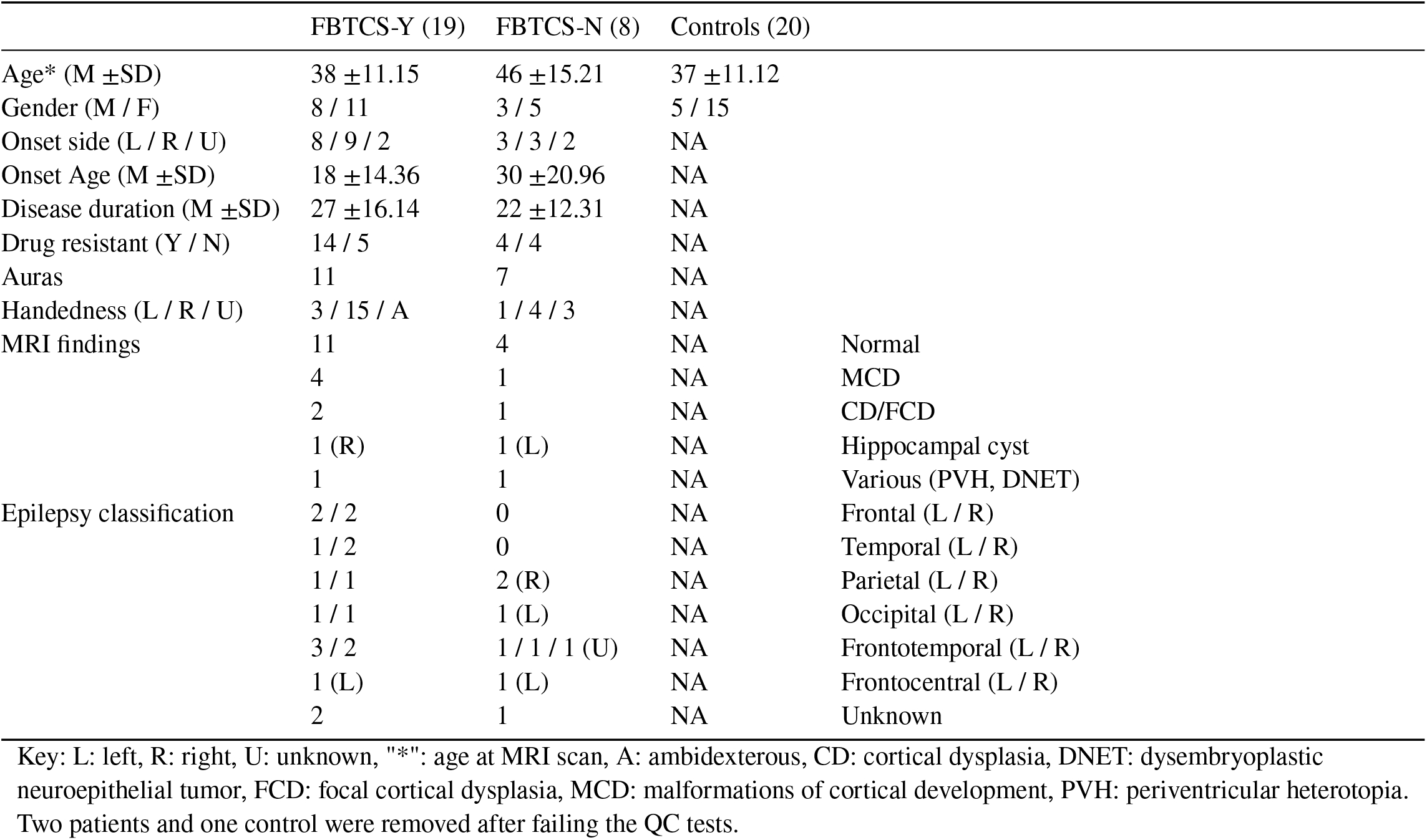
Characteristics of patients and control participants.

### 2.2 All patients versus Controls

Individual analysis of covariance (ANCOVA) tests were conducted to examine the between-group differences in the two grouping conditions (“All patients” versus controls; and FBTCS-Y and FBTCS-N versus controls). The tracts of interest were: Thalamus - Prefrontal (TPREF), Premotor (TPREM), Precentral (TPREC), Postcentral (TPOSTC), Parietal (TPAR), and Occipital (TOCC); Striato - Fronto-Orbital (STFO), Prefrontal (STPREF), Premotor (STPREM), Precentral (STPREC), Postcentral (STPOSTC), Parietal (STPAR), and Occipital (STOCC); Anterior thalamic radiation (ATR), Superior thalamic radiation (STR), Corticospinal (CST), Fronto pontine (FPT), Parieto-occipital pontine (POPT), Inferior cerebellar peduncle (ICP), Middle cerebellar peduncle (MCP), Superior cerebellar peduncle (SCP), Inferior fronto-occipital fascicle (IFO), Uncinate fascicle (UF), Commissure anterior (CA), and Corpus callosum (CC).

Only the tracts that demonstrated a significant difference in mean TW-TM are reported here (*p* values extracted from the pairwise comparisons), all other results are reported in the Supplementary Tables. Where more than five tracts showed significant between-group differences for a given TW-TM, the *p* value is reported as *p* < 0.05, with exact *p* values provided in the Supplementary Tables.

The whole-tract, average TW-ADC was higher in the “All patients” group compared to controls, for 16 out of 24 tracts (*p* < 0.05). The UF tract had a lower average TW-FA in the “All patients” group compared to controls (*p* = 0.023). Nineteen tracts had higher average TW-RD in the “All patients” group compared to controls(*p* < 0.05). The MCP and SCP had higher average TW-AD in “All patients” compared to controls (*p* = 0.033*andp* = 0.032 respectively). Figure 1 (a) shows the mean TW-TM of the “All patients” group compared to controls. When group differences were tested in each hemisphere, the previously observed significant whole-tract differences were not preserved in both hemispheres (shown in Figure 1, b). All mean differences, exact *p* values and confidence intervals for the “All patients” versus controls comparison are reported in Supplementary Tables 1 and 2.

**FIGURE 1.**
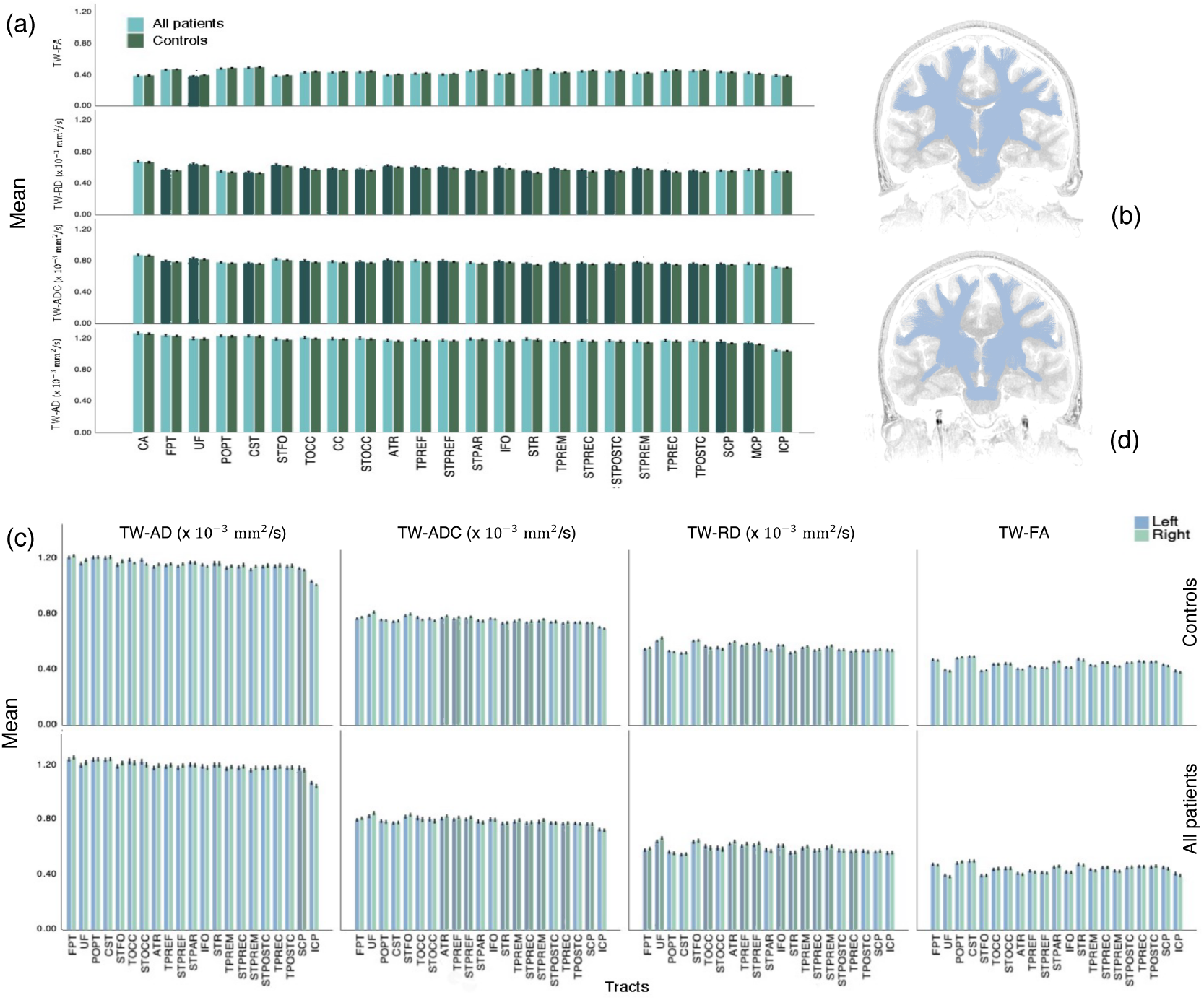
Bar graphs of mean TW-TM differences between All patients and controls. The mean differences in the wholetract TW-TM are shown in (a); error bars indicate the 95% confidence intervals (CI). The darker bars indicate the tracts where the difference between the two groups was significant (*p* < 0.05); (b) shows a two-dimensional visual representation of the significant tracts. The mean difference in the TW-TM for the left and right hemispheres are shown in (c), error bars indicate the 95% CIs. Here, the darker bars denote tracts where the difference between the two groups was significant in both hemispheres (*p* < 0.05); (d) visually represents the lower number of left and right tracts displaying significant, between-group differences compared to the whole-tract analysis.

### 2.3 FBTCS-Y, FBTCS-N versus Controls

Group differences were observed in a number of TW-TM in the whole-tract comparison. Here, we report the *p* value from the post hoc tests which revealed that overall, the significant difference between the FBTCS-N and control groups drove the main effects observed in ANCOVAs (see Supplementary Table 3). The average TW-ADC was higher for the FBTCS-N group compared to controls for 18 out of 24 tracts (*p* < 0.05), and in the average TW-FA of the CC (*p* = 0.043), TPREF (*p* = 0.048), and TPREM tracts(*p* = 0.031). The average TW-RD was higher for the FBTCS-N group compared to controls for 18 out of 24 tracts (*p* < 0.05). The FBTCS-N group also had higher TW-AD in the IFO (*p* = 0.030), STOCC (*p* = 0.018) and TOCC (*p* = 0.026) tracts. Interestingly, the FBTCS-N group had significantly higher TW-ADC and TW-AD in several tracts compared to the FBTCS-Y group (TW-ADC - IFO: *p* = 0.044, STOCC: *p* = 0.032, TOCC: *p* = 0.042; TW-AD - IFO: *p* = 0.049, STOCC: *p* = 0.033). The FBTCS-Y group also had significantly higher TW-AD than the control group in the MCP (*p* = 0.042). Figure 2 (a) shows the mean TW-TM for the three groups. When group differences were tested in each hemisphere, the previously observed significant whole-tract differences were not preserved in both hemispheres.

**FIGURE 2.**
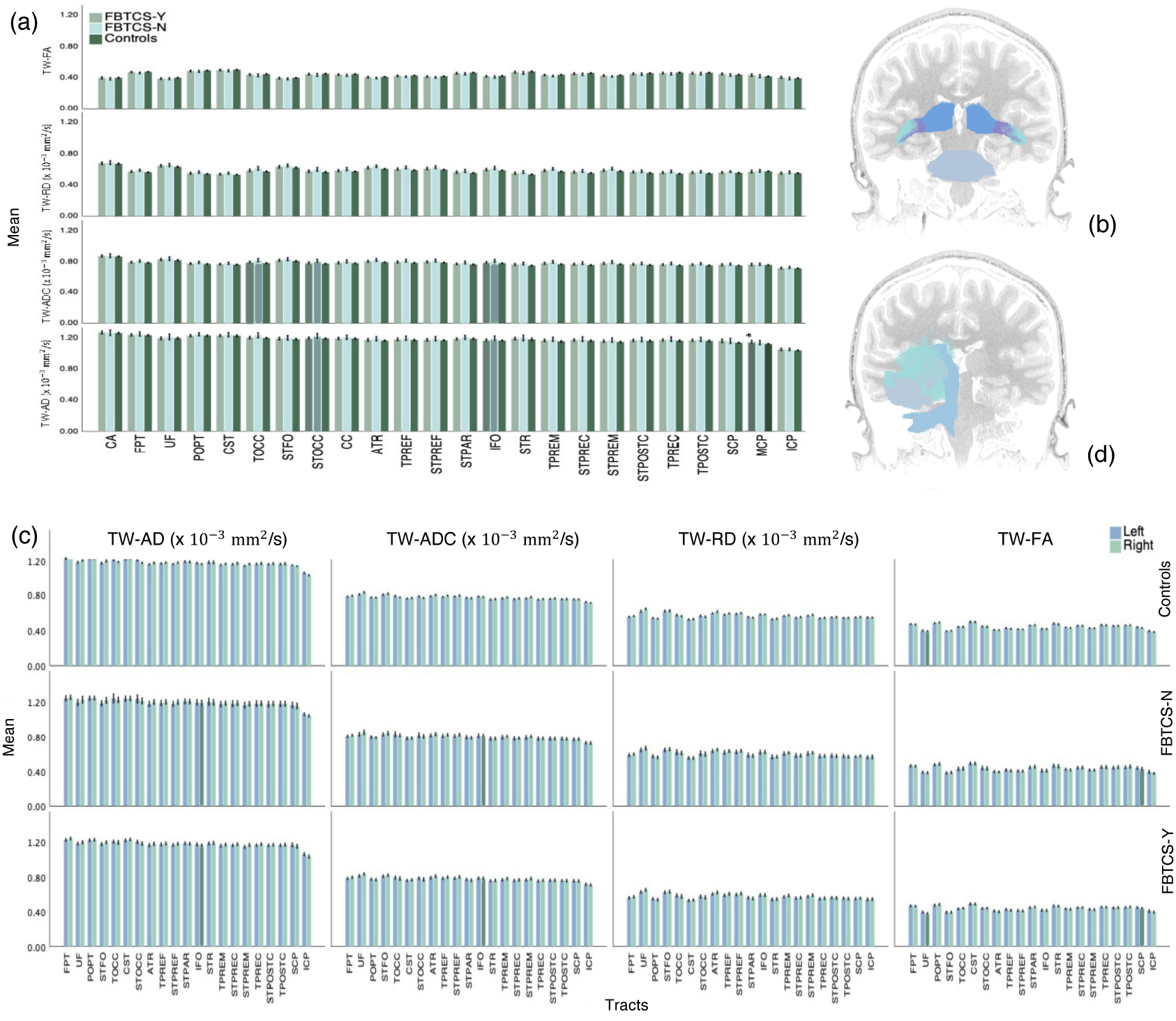
Bar graphs of TW-TM differences between the FBTCS-Y, FBTCS-N and control groups. The mean TW-TM differences in the whole-tracts shown in (a), error bars indicate the 95% CIs. The darker bars indicate the t racts where the difference between FBTCS-Y and FBTCS-N groups was significant (*p* < 0.05); (b) shows a visual representation of the tracts that displayed significant between-group differences (blue: TOCC, purple: STOCC, green: IFO, grey-blue: MCP). The mean TW-TM differences in the left and right hemispheres are shown in (c). Here, the darker bars highlight the tracts where a significant mean difference was observed between the FBTCS-Y and FBTCS-N group, or between the FBTCS-Y group and controls (indicated by “*” on MCP tract) in the left or right hemisphere; (d) shows a visual representation of the tracts that displayed significant mean differences (green: IFO, grey-blue: UF, light blue: SCP).

Importantly, compared to the FBTCS-Y group, the significantly higher TW-ADC and TW-AD of the IFO tract in the FBTCS-group was retained only in the right side (*p* = 0.047 and *p* = 0.028 respectively, shown in Figure 2 (c)). All mean differences, act values, confidence intervals and main effects for the ANCOVAs between the FBTCS-Y, FBTCS-N and controls are provided in Supplementary Tables 3 and 4.

## 3 DISCUSSION

Distinct from previous works, this study implemented an automated, track-weighted tensor imaging pipeline to examine the microstructural white matter changes in patients with and without FBTCS compared to controls.

Our study revealed unexpected, marked alterations in subcortical tracts considered critical to seizure genesis. Our findings indicated that the type of white matter disruption (i.e., TW-AD) in combination with the disrupted region, may be relevant to the seizure semiology observed in FBTCS. Our results show that the region-specific alterations are likely a function of seizure pathology rather than whole-brain differences. The ensuing discussion focuses solely on the compelling results that could yield clinical utility; readers may refer to the Supplementary tables to further examine other results.

One of the remarkable findings was the significant difference (in TW-ADC, TW-AD) between the two patient subgroups in the IFO and STOCC and the TW-ADC of the TOCC tract. In patients with mesio-TLE, stereotactic EEG (SEEG) has shown the role of thalamocortical connections in seizure termination^24^. However, altered thalamic functional profiles have also been proposed as imaging biomarkers of active secondary generalisation^25^. Specifically, the thalamocortical circuit has been implicated as a critical mechanism for the genesis of FBTCS^26^ by engagement of the striatum and CC.

The inhibitory role of the BG has been shown in TLE yet remains unclear in FBTCS. Mounting research using SEEG, DBS, and EEG/fMRI suggest the physiological rhythms in the BG as a “pacemaker” for ictal discharge throughout adjacent regions^12^. SEEG demonstrated cortico-striatal synchronisation^27^, implicating the changing synchronisation as a mechanism to control the duration of abnormal oscillations within the striatal-thalamo-cortical loop and for potential termination. DBS studies show that regulation of BG activity via thalamostriatal projections may mediate generalised seizures^28^. Combined EEG/fMRI measurements showed the thalamocortical-striatal network could be involved in activation (thalamocortical), deactivation (striatal) and consequently termination of cortical discharge^29^. The cortico-striato-thalamo-cerebellar network has been implicated as a prominent feature of FBTCS via increased connectivity of structural covariance in the striatum and thalamus^30^. Here, we identified specific tracts (IFO, STOCC, TOCC) that may represent a predictive biomarker to differentiate between individuals who develop FBTCS versus those who do not. The increased TW-ADC in conjunction with the increased TW-AD could indicate white matter disruption significant enough to inhibit propagation of ictal discharge. This finding could guide researchers in investigating the specific circuits and networks involved in FBTCS.

Secondly, the finding of a significant difference between the FBTCS-N and control groups in the TW-ADC, TW-FA and TW-RD of the TPREF and TPREM tracts might be explained by the gatekeeping function of the subcortical structures. The white matter disruption in those tracts may induce changes to their primary function in seizure activity, acting as a protective mechanism that inhibits seizure propagation. Increased degradation of the fibre bundles that form the seizure propagation pathways may diminish the likelihood of ictal discharge traversing to the contralateral hemisphere. However, for those with less altered TPREF and TPREM tracts, such as in the FBTCS-Y group, the white matter fibre bundles may bear a closer resemblance to the control participants, and ictal discharge events could gain enough momentum to override the inhibitory mechanisms and cross to the contralateral hemisphere.

Alternatively, the white matter disruption in the FBTCS-N group may result from elevated BG activity during seizure inhibition. Extensive overworking of the pathways in the subcortical regions could manifest as axonal damage. In TLE, patterns of electrophysiological discharge propagation are purportedly bolstered by the structural white matter pathways, which reinforce ictal propagation^31^, suggesting that white matter damage may halt the natural progression of ictal discharge.

Third, considering that the overall significant differences were observed between the FBTCS-N group and controls, the significantly higher TW-AD of the MCP in the FBTCS-Y group is noteworthy. Anatomically, the MCP crosses the two hemispheres, connecting the cerebellum to the pons, and is composed entirely of centripetal fibres, i.e. incoming fibres. Previous work has shown the role of the MCP in generalised tonic-clonic seizures^32^, potentiating it as a tract whereby microstructural damage may impart susceptibility to the development of a generalised seizure.

Although increased AD has been shown to result from white matter maturation, all groups were age-matched, alluding to the possibility that the higher TW-AD in the MCP of the FBTCS-Y group may be due to distinct FBTCS mechanisms. Since increased AD can also imply better organisation of fibre structure, the TW-AD differences between the FBTCS-Y and FBTCS-N groups in the cerebellar region could be a feature of the FBTCS brain whereby those with FBTCS-Y have better fibre connections and the ability to sustain the intrahemispheric flow of ictal discharge. Conversely, our FBTCS-N group may have more damaged or poor fibre connections, evidenced by the overall higher TW-ADC and TW-RD in the thalamus, striatal and CC tracts compared to the FBTCS-Y and control groups. The MCP tract could present a compelling region of interest in post-surgical imaging, and future works could investigate this further. Lastly, though the ICP is part of the peduncles and is anatomically adjacent to the SCP and MCP, it is not functionally important in inhibition nor information relay, which may explain the absence of significant difference in this tract between the groups. Our findings promote the possibility that propagation of ictal discharge from the epileptogenic zone to the contralateral hemisphere could be inhibited by damage to critical information relay and anatomically relevant tracts.

Finally, there was no significant relationship between lesion presence and seizure onset side, or lesion presence and FBTCS, reinforcing the conjecture that the region-specific differences between the patient subgroups were due to premorbid connections rather than the effects of seizure injury.

Imaging studies of patients with epilepsy are traditionally limited by small sample sizes^22,33,21,23^, primarily due to challenges in recruitment. Our relatively small sample size was a limitation in our study, which prevented further analysis of the striatal and thalamus tracts. Nevertheless, our study emphasises that specific tracts may play a role in FBTCS, reinforcing the value of structural imaging in demystifying the mechanisms involved in FBTCS. Though generalised seizures may appear similar in semiology to FBTCS, specific pathways and networks may be involved in FBTCS once the bi-hemispheric ictal propagation begins and is an essential subject for further research.

In summary, we used advanced diffusion MRI to show region-specific microstructural alterations in patients with focal epilepsy compared to control participants. Our findings provide mechanistic insights into how structural changes may impact the functional role of the thalamic and striatal regions in individuals with FBCTS. We highlight specific tracts (IFO, STOCC, TPREF and TPREM) that may be involved in FBTCS. Our results lay the foundation for a better understanding of seizure propagation in FBCTS, and offer potential biomarkers that can help explain disease progression and aid treatment.

## 4 METHODS

### 4.1 Participants and Data

Twenty-seven adults with focal epilepsy were recruited from the Comprehensive Epilepsy Centre at the Royal Prince Alfred Hospital (RPAH, Sydney, Australia); and MRI was performed at the Brain and Mind Centre (Sydney, Australia). Inclusion criteria were adults diagnosed with focal epilepsy, aged 18-60, presenting without surgery, and with or without a cortical brain lesion, who were willing and able to comply with the study procedures for the duration of their participation. Exclusion criteria were pregnant women and individuals with intellectual disabilities. The 20 controls were neurologically normal individuals. Written informed consent was obtained from all participants before study participation. Ethical approval was obtained from the RPAH and the University of Sydney ethics bodies (RPAH approval ID: X14-0347; University of Sydney approval ID: HREC/14/RPAH/467) and the study conducted in accordance with guidelines set forth by those entities.

#### 4.1.1 Epilepsy patient groups

The participants with focal epilepsy were placed into the following groups:

1. “All patients”: The entire cohort of patients; all diagnosed with focal epilepsy.
2. “FBTCS-Y”: In this subgroup, patients were defined as having frequent (more than two per year) or infrequent (one per year) large, homolateral and simultaneous FBTCS. The FBTCS may have occurred during observation at the RPAH Epilepsy Centre or reported by the participant as occurring elsewhere.
3. “FBTCS-N”: In this subgroup, patients had never experienced FBTCS.

### 4.2 Image acquisition

All scans were acquired on the same GE Discovery™ MR750 3T scanner (GE Medical Systems, Milwaukee, WI). For each participant, the following sequences were acquired: Pre-contrast 3D high-resolution T1-weighted image (0.7mm isotropic) using fast spoiled gradient echo (SPGR) with magnetisation-prepared inversion recovery pulse (TE/TI/TR=2.8/900/7.1ms, flip angle=12); and axial diffusion-weighted imaging (2mm isotropic, TE/TR=85/8325ms) with a uniform gradient loading (*b*=1000s/mm^2^) in 64 directions and 2 *b* 0s. An additional *b*0 image with reversed phase-encoding was also acquired for distortion correction^34^.

### 4.3 Image preprocessing

The T1 images were processed using a modified version of Freesurfer’s recon-all (v6.0)^35^, alongside an in-house skull-stripping tool (Sydney Neuroimaging Analysis Centre). Each subject was inspected, and minor segmentation errors were manually corrected. A 5 tissue-type (5TT) image was generated using MRtrix3^36^. The T1 image was registered to the mean *b*0 image; the warp was used to register the 5TT image to the diffusion image.

Diffusion image processing was conducted using MRtrix3^36^. The diffusion pre-processing included motion and distortion correction^34,37^, bias correction using ANTS^38^, and resizing to voxel size 1 mm isotropic. The *dhollander* algorithm^39^ was used to estimate the response functions of the white matter, grey matter, and cerebral spinal fluid, from which constrained spherical deconvolution was used to estimate the fibre orientation distributions using MRtrix3Tissue^36^. The intensity of the white matter fibre orientation distributions was normalised^36^, and used for anatomically constrained whole-brain tractography^40^. The tractography settings were: 15 million tracks were generated, iFOD2 probabilistic fibre tracking^41^, dynamic seeding^42^, maximum length 300 mm, backtrack selected and crop at grey-matter-white-matter interface selected. For quantitative analysis, the corresponding weight for each streamline in the tractogram was derived using SIFT2^42^; the streamline weights and tractogram were used to generate a track-density image (TDI)^43^.

#### Track-weighted tensor-based measurements

The pre-processed diffusion image was used to calculate the diffusion tensor. By resolving the tensor into its primary, secondary and tertiary directions of diffusion, the following tensor-based metrics can be calculated: apparent diffusion coefficient (ADC, the average of the three directions of diffusion), fractional anisotropy (FA, degree of anisotropy ranging from 0 to 1, whereby 0 indicates isotropic diffusion and 1 indicates diffusion exclusively along a single axis), axial diffusivity (AD, the primary direction of diffusion, representative of diffusion parallel to axonal fibres) and radial diffusivity (RD, the average of the secondary and tertiary directions of diffusion, representing diffusion perpendicular to axonal fibres)^44^. The tensor metrics (ADC, FA, AD, and RD) were estimated. Next, the tracks (and their weights) were used to calculate the track-weighted (TW) tensor-based metrics (i.e., TW-ADC, TW-FA, TW-AD, TW-RD)^45^, known to improve reproducibility and variability (compared to standalone tensor-based metrics)^46^. The following settings were used to generate TW images: Gaussian statistic, full-width-half-maximum of 40 and voxel size of 0.2mm.

#### Measuring tract-specific, track-weighted tensor-based metrics

Tractseg^47^, a superior automated tract segmentation model, was applied to the resized diffusion images to obtain data-driven, subject-specific segmentations of the selected tracts. A schematic of the image analysis pipeline is shown in Figure 3. Two brain imaging specialists (CW and AD) conducted a quality control check by visually inspecting each tract for consistency and anatomical correctness. The right TPAR tract failed the quality control check in 10 participants; therefore the whole tract was removed from the analysis. Twenty-four tracts were included in the final statistical analysis.

**FIGURE 3.**
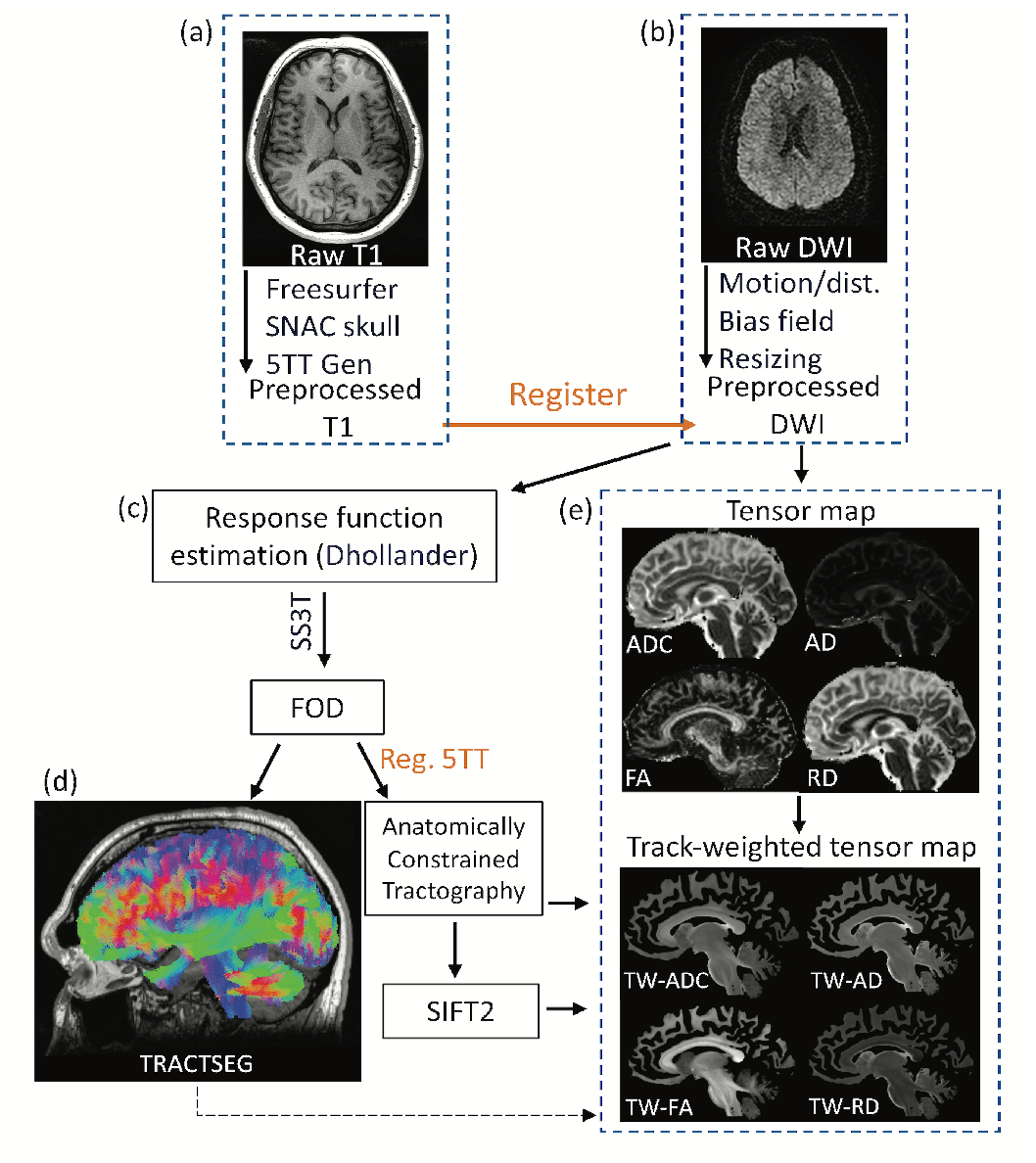
Schematic overview of imaging analysis pipeline. T1 (a) and diffusion MRIs (b) were preprocessed as described in Methods section 4.3. Next, further diffusion image processing and tractography was conducted (c) and tract segmentation performed (d). Finally, track-weighted tensor maps were produced, and the resulting metrics were derived (e).

### 4.4 Statistical analysis

#### Demographics

The chi-square test was used to investigate the presence of the following associations: lesion presence and seizure onset side; and lesion presence and FBTCS; and gender and age differences between the “All patients” and control groups, and the FBTCS-Y, FBTCS-N and control groups.

#### Group comparisons: All patients versus controls

To examine whole-tract differences between groups, the weighted average of the left and right side of each tract was calculated using the equation [WeightedAverage-TW-TM = ((*L*_TW*−*TM_ × *L*_count_) + (*R*_TW*−*TM_ × *R*_count_))⁄(*L*_count_ + *R*_count_)] (TW-TM: track-weighted tensor metric i.e. TW-ADC; count: tract count). Univariate ANCOVAs (which are robust to multiple comparisons including covariates^48^) were used to examine the whole-tract differences for each TW-TM (i.e. TW-ADC, TW-FA) between the “All patients” and control groups. A separate set of ANCOVAs was employed to investigate group differences in TW-TM in a given hemisphere. Here, the TW-TM for a given tract from a given hemisphere was compared between the “All patients” and control groups.

#### Group comparisons: FBTCS-Y, FBTCS-N versus Controls

ANCOVAs were used to investigate differences between the patient subgroups (FBTCS-Y, FBTCS-N) and control participants. Again, the weighted average of each tract’s left and right sides was computed (as described above) to investigate whole-tract differences in TW-TM between the three groups. As with the previous grouping condition, ANCOVAs were then used to investigate group differences in a given hemisphere. Here, the TW-TM for a given tract from a given hemisphere was compared between the three groups.

To account for age effects on diffusivity measures^49^, age was included as a covariate in all the ANCOVAs. The threshold for statistical significance was set at *p* = 0.05 for all ANCOVAs. In each group comparison, the mean difference, *p* value and confidence intervals for each tract in each TW*−*TM were taken from the Bonferroni adjusted pairwise comparisons generated from the estimated marginal means, which is robust against unbalanced groups and multiple comparisons. All statistical analyses were conducted in SPSS v28.

## Data Availability

The data that support the findings of this study are available from Royal Prince Alfred Hospital/The University of Sydney. However, restrictions apply to the availability of these data, which were used under license/ethical approval for the current study, and so are not publicly available.

## 5 ACKNOWLEDGEMENTS

The authors acknowledge all staff at the Comprehensive Epilepsy Centre at the RPAH, particularly Mrs Maricar Senturias (RN/ACNC Epilepsy), who assisted with patient recruitment. The authors acknowledge the radiology staff at i-MED Radiology for their assistance with obtaining the MRI data. The authors acknowledge the research funding support from UCB Australia Pty Ltd. CM acknowledges scholarship support from the Nerve Research Foundation, University of Sydney. AD acknowledges funding from St. Vincent’s Hospital. OK acknowledges the partial support provided by The University of Sydney through a SOAR Fellowship and Microsoft’s partial support through a Microsoft AI for Accessibility grant. CW acknowledges research funding from the Nerve Research Foundation, University of Sydney.

## 7 DATA AVAILABILITY

The datasets generated during and/or analysed during the current study are not publicly available because they are RPAH patients, and access is currently only for authorised individuals named on the approved ethics application. However, de-identified data can be made available from the corresponding author on reasonable request, subject to approval from the relevant governing ethics entities at the RPAH and The University of Sydney.

## 8 AUTHOR CONTRIBUTIONS

CM: Conceptualisation, methodology, data curation, imaging analysis, statistical analysis, manuscript writing, review and editing. AD: Imaging and diffusion analysis pipeline implementation, data curation and analysis, manuscript revision. RZ: Imaging pipeline-Data acquisition and Tractseg implementation. MB and OK: Conceptualisation and manuscript revision. AN: Clinical data acquisition, clinical advisory, conceptualisation, methodology, manuscript revision. CW: Conceptualisation, methodology, data curation, imaging analysis, statistical analysis, manuscript revision.

