## Supplementary Table 1 for "White matter alterations in focal to bilateral tonic-clonic seizures" (from another version of this preprint: version 1, 2021-10-26)

**Supplementary Table 1.** Whole-tract, mean differences between the “all patients” and control groups by track-weighted tensor metric.

| | TW-ADC | | | | TW-FA | | | | TW-RD | | | | TW-AD | | | |
|---|---|---|---|---|---|---|---|---|---|---|---|---|---|---|---|---|
| Tracts | Mean Difference | *p* | 95% CI - Lower | 95% CI - Upper | Mean Difference | *p* | 95% CI - Lower | 95% CI - Upper | Mean Difference | *p* | 95% CI - Lower | 95% CI - Upper | Mean Difference | *p* | 95% CI - Lower | 95% CI - Upper |
| Anterior thalamic radiation (ATR) | 1.687E-5* | 0.013 | 3.685E-06 | 3.005E-05 | -0.007 | 0.114 | -0.016 | 0.002 | 1.826E-5* | 0.008 | 5.150E-06 | 3.137E-05 | 1.408E-05 | 0.103 | -2.974E-06 | 3.114E-05 |
| Superior thalamic radiation (STR) | 1.727E-5* | 0.010 | 4.427E-06 | 3.012E-05 | -0.010 | 0.120 | -0.024 | 0.003 | 1.956E-5* | 0.008 | 5.312E-06 | 3.381E-05 | 1.270E-05 | 0.207 | -7.331E-06 | 3.273E-05 |
| Corticospinal (CST) | 1.106E-5* | 0.044 | 3.328E-07 | 2.178E-05 | -0.007 | 0.187 | -0.018 | 0.004 | 1.239E-5* | 0.043 | 4.347E-07 | 2.435E-05 | 8.391E-06 | 0.300 | -7.769E-06 | 2.455E-05 |
| Fronto pontine (FPT) | 1.277E-5* | 0.035 | 9.409E-07 | 2.459E-05 | -0.006 | 0.155 | -0.016 | 0.003 | 1.416E-5* | 0.026 | 1.753E-06 | 2.656E-05 | 9.974E-06 | 0.203 | -5.595E-06 | 2.554E-05 |
| Parieto-occipital pontine (POPT) | 1.003E-05 | 0.078 | -1.175E-06 | 2.123E-05 | -0.008 | 0.144 | -0.018 | 0.003 | 1.232E-05 | 0.052 | -1.193E-07 | 2.476E-05 | 5.451E-06 | 0.463 | -9.428E-06 | 2.033E-05 |
| Inferior cerebellar peduncle (ICP) | 6.203E-06 | 0.268 | -4.957E-06 | 1.736E-05 | 0.004 | 0.502 | -0.008 | 0.017 | 3.340E-06 | 0.604 | -9.584E-06 | 1.626E-05 | 1.193E-05 | 0.100 | -2.387E-06 | 2.625E-05 |
| Middle cerebellar peduncle (MCP) | 6.455E-06 | 0.318 | -6.457E-06 | 1.937E-05 | 0.011 | 0.130 | -0.003 | 0.026 | -1.968E-07 | 0.979 | -1.519E-05 | 1.479E-05 | 1.976E-5* | 0.033 | 1.668E-06 | 3.785E-05 |
| Superior cerebellar peduncle (SCP) | 1.234E-5* | 0.026 | 1.528E-06 | 2.316E-05 | 0.004 | 0.441 | -0.007 | 0.016 | 7.848E-06 | 0.142 | -2.731E-06 | 1.843E-05 | 2.1331E-5* | 0.032 | 1.963E-06 | 4.070E-05 |
| Inferior fronto occipital (IFO) | 1.446E-5* | 0.042 | 5.233E-07 | 2.839E-05 | -0.008 | 0.154 | -0.018 | 0.003 | 1.631E-5* | 0.031 | 1.557E-06 | 3.105E-05 | 1.076E-05 | 0.190 | -5.546E-06 | 2.706E-05 |
| ST_ Fronto orbital (STFO) | 1.374E-05 | 0.057 | -4.027E-07 | 2.789E-05 | -0.007 | 0.158 | -0.017 | 0.003 | 1.514E-5* | 0.043 | 5.312E-07 | 2.974E-05 | 1.095E-05 | 0.211 | -6.455E-06 | 2.836E-05 |
| ST_Occipital (STOCC) | 1.554E-5* | 0.046 | 2.660E-07 | 3.082E-05 | -0.007 | 0.247 | -0.019 | 0.005 | 1.706E-5* | 0.044 | 5.097E-07 | 3.362E-05 | 1.250E-05 | 0.162 | -5.226E-06 | 3.022E-05 |
| ST_Parietal (STPAR) | 1.166E-05 | 0.077 | -1.335E-06 | 2.466E-05 | -0.010 | 0.093 | -0.021 | 0.002 | 1.460E-5* | 0.045 | 3.390E-07 | 2.886E-05 | 5.784E-06 | 0.455 | -9.705E-06 | 2.127E-05 |
| ST_Postcentral (STPOSTC) | 1.344E-5* | 0.028 | 1.505E-06 | 2.537E-05 | -0.009 | 0.124 | -0.021 | 0.003 | 1.498E-5* | 0.026 | 1.887E-06 | 2.807E-05 | 1.036E-05 | 0.216 | -6.291E-06 | 2.701E-05 |
| ST_Precentral (STPREC) | 1.599E-5* | 0.014 | 3.384E-06 | 2.860E-05 | -0.010 | 0.081 | -0.021 | 0.001 | 1.754E-5* | 0.013 | 3.970E-06 | 3.112E-05 | 1.288E-05 | 0.122 | -3.576E-06 | 2.934E-05 |
| ST_Prefrontal (STPREF) | 1.532E-5* | 0.026 | 1.948E-06 | 2.869E-05 | -0.008 | 0.094 | -0.017 | 0.001 | 1.691E-5* | 0.017 | 3.183E-06 | 3.064E-05 | 1.213E-05 | 0.134 | -3.882E-06 | 2.815E-05 |
| ST_Premotor (STPREM) | 1.575E-5* | 0.024 | 2.218E-06 | 2.928E-05 | -0.007 | 0.126 | -0.017 | 0.002 | 1.638E-5* | 0.020 | 2.679E-06 | 3.008E-05 | 1.448E-05 | 0.088 | -2.263E-06 | 3.123E-05 |
| T_Occipital (TOCC) | 1.790E-5* | 0.027 | 2.167E-06 | 3.363E-05 | -0.008 | 0.166 | -0.020 | 0.004 | 1.989E-5* | 0.022 | 3.064E-06 | 3.672E-05 | 1.392E-05 | 0.131 | -4.334E-06 | 3.217E-05 |
| T_Postcentral (TPOSTC) | 1.360E-5* | 0.019 | 2.387E-06 | 2.480E-05 | -0.009 | 0.160 | -0.021 | 0.004 | 1.487E-5* | 0.022 | 2.215E-06 | 2.753E-05 | 1.104E-05 | 0.192 | -5.790E-06 | 2.787E-05 |
| T_Precentral (TPREC) | 1.688E-5* | 0.008 | 4.623E-06 | 2.913E-05 | -0.010 | 0.075 | -0.022 | 0.001 | 1.864E-5* | 0.008 | 5.213E-06 | 3.207E-05 | 1.335E-05 | 0.112 | -3.231E-06 | 2.993E-05 |
| T_Prefrontal (TPREF) | 1.686E-5* | 0.014 | 3.647E-06 | 3.008E-05 | -0.008 | 0.091 | -0.017 | 0.001 | 1.839E-5* | 0.009 | 4.870E-06 | 3.191E-05 | 1.381E-05 | 0.094 | -2.460E-06 | 3.008E-05 |
| T_Premotor (TPREM) | 1.728E-5* | 0.009 | 4.463E-06 | 3.009E-05 | -0.007 | 0.143 | -0.017 | 0.003 | 1.773E-5* | 0.009 | 4.750E-06 | 3.071E-05 | 1.637E-05 | 0.055 | -4.007E-07 | 3.314E-05 |
| Uncinate fascicle (UF) | 1.324E-05 | 0.066 | -9.223E-07 | 2.741E-05 | -.012* | 0.023 | -0.022 | -0.002 | 1.671E-5* | 0.020 | 2.729E-06 | 3.070E-05 | 6.286E-06 | 0.509 | -1.277E-05 | 2.534E-05 |
| Commissure anterior (CA) | 8.247E-06 | 0.253 | -6.128E-06 | 2.262E-05 | -0.004 | 0.552 | -0.018 | 0.010 | 8.290E-06 | 0.309 | -7.986E-06 | 2.457E-05 | 8.160E-06 | 0.416 | -1.191E-05 | 2.823E-05 |
| Corpus callosum (CC) | 1.287E-05 | 0.057 | -3.895E-07 | 2.614E-05 | -0.009 | 0.065 | -0.020 | 0.001 | 1.584E-5* | 0.030 | 1.613E-06 | 3.007E-05 | 6.936E-06 | 0.349 | -7.867E-06 | 2.174E-05 |

Key: Mean difference: the mean difference of the "all patients" group minus the control group, based on the estimated marginal means of the univariate ANCOVA. Positive mean difference values indicate the "all patients" group had a higher average TW-TM than the control group, negative values indicate the control group had a higher TW-TM than "all patients". "*p*" presents the significance of the mean difference. * a significant difference was observed in the estimated marginal means univariate ANCOVA.
