## Supplementary Table 2 for "White matter alterations in focal to bilateral tonic-clonic seizures" (from another version of this preprint: version 1, 2021-10-26)

**Supplementary Table 2.** Pairwise comparisons of the track-weighted, mean differences between the "all patients" and control groups for each left and right side of a given tract.

**TW-ADC**

| Tract | Left: Mean Difference | Left: *p* | Left: 95% CI - Lower | Left: 95% CI - Upper | Right: Mean Difference | Right: *p* | Right: 95% CI - Lower | Right: 95% CI - Upper |
|---|---|---|---|---|---|---|---|---|
| Anterior thalamic radiation (ATR) | 1.552E-5* | 0.022 | 2.311E-06 | 2.874E-05 | 1.832E-5* | 0.009 | 4.844E-06 | 3.180E-05 |
| Superior thalamic radiation (STR) | 1.828E-5* | 0.008 | 5.085E-06 | 3.147E-05 | 1.648E-5* | 0.018 | 2.946E-06 | 3.001E-05 |
| Corticospinal (CST) | 1.143E-5* | 0.043 | 3.662E-07 | 2.249E-05 | 1.074E-05 | 0.059 | -4.379E-07 | 2.191E-05 |
| Fronto pontine (FPT) | 1.189E-05 | 0.053 | -1.414E-07 | 2.392E-05 | 1.370E-5* | 0.025 | 1.828E-06 | 2.556E-05 |
| Parieto-occipital pontine (POPT) | 1.122E-05 | 0.055 | -2.591E-07 | 2.270E-05 | 9.192E-06 | 0.142 | -3.223E-06 | 2.161E-05 |
| Inferior cerebellar peduncle (ICP) | 4.252E-06 | 0.438 | -6.714E-06 | 1.522E-05 | 8.313E-06 | 0.185 | -4.149E-06 | 2.077E-05 |
| Superior cerebellar peduncle (SCP) | 1.248E-5* | 0.025 | 1.662E-06 | 2.331E-05 | 1.231E-5* | 0.034 | 9.515E-07 | 2.367E-05 |
| Inferior fronto occipital (IFO) | 1.405E-05 | 0.054 | -2.611E-07 | 2.835E-05 | 1.534E-5* | 0.042 | 5.870E-07 | 3.009E-05 |
| ST_ Fronto orbital (STFO) | 1.256E-05 | 0.078 | -1.485E-06 | 2.662E-05 | 1.541E-5* | 0.044 | 4.107E-07 | 3.040E-05 |
| ST_Occipital (STOCC) | 1.462E-05 | 0.091 | -2.437E-06 | 3.167E-05 | 1.791E-5* | 0.035 | 1.290E-06 | 3.454E-05 |
| ST_Parietal (STPAR) | 1.272E-05 | 0.058 | -4.549E-07 | 2.590E-05 | 1.099E-05 | 0.122 | -3.049E-06 | 2.502E-05 |
| ST_Postcentral (STPOSTC) | 1.555E-5* | 0.011 | 3.812E-06 | 2.728E-05 | 1.112E-05 | 0.100 | -2.208E-06 | 2.445E-05 |
| ST_Precentral (STPREC) | 1.738E-5* | 0.009 | 4.507E-06 | 3.026E-05 | 1.450E-5* | 0.034 | 1.125E-06 | 2.789E-05 |
| ST_Prefrontal (STPREF) | 1.428E-5* | 0.035 | 1.019E-06 | 2.755E-05 | 1.647E-5* | 0.020 | 2.741E-06 | 3.019E-05 |
| ST_Premotor (STPREM) | 1.580E-5* | 0.027 | 1.885E-06 | 2.972E-05 | 1.574E-5* | 0.025 | 2.067E-06 | 2.942E-05 |
| T_Occipital (TOCC) | 1.697E-05 | 0.072 | -1.581E-06 | 3.552E-05 | 2.075E-5* | 0.020 | 3.470E-06 | 3.802E-05 |
| T_Postcentral (TPOSTC) | 1.551E-5* | 0.007 | 4.434E-06 | 2.659E-05 | 1.165E-05 | 0.071 | -1.057E-06 | 2.436E-05 |
| T_Precentral (TPREC) | 1.756E-5* | 0.007 | 5.042E-06 | 3.008E-05 | 1.624E-5* | 0.016 | 3.252E-06 | 2.922E-05 |
| T_Prefrontal (TPREF) | 1.589E-5* | 0.020 | 2.660E-06 | 2.911E-05 | 1.795E-5* | 0.011 | 4.437E-06 | 3.146E-05 |
| T_Premotor (TPREM) | 1.670E-5* | 0.016 | 3.276E-06 | 3.012E-05 | 1.807E-5* | 0.007 | 5.304E-06 | 3.084E-05 |
| Uncinate fascicle (UF) | 1.386E-5* | 0.045 | 3.462E-07 | 2.738E-05 | 1.370E-05 | 0.081 | -1.770E-06 | 2.918E-05 |

**TW-FA**

| Tract | Left: Mean Difference | Left: *p* | Left: 95% CI - Lower | Left: 95% CI - Upper | Right: Mean Difference | Right: *p* | Right: 95% CI - Lower | Right: 95% CI - Upper |
|---|---|---|---|---|---|---|---|---|
| Anterior thalamic radiation (ATR) | -0.006 | 0.246 | -0.015 | 0.004 | -.009* | 0.044 | -0.018 | 0.000 |
| Superior thalamic radiation (STR) | -0.013 | 0.066 | -0.026 | 0.001 | -0.008 | 0.218 | -0.022 | 0.005 |
| Corticospinal (CST) | -0.007 | 0.211 | -0.018 | 0.004 | -0.007 | 0.183 | -0.018 | 0.003 |
| Fronto pontine (FPT) | -0.006 | 0.175 | -0.015 | 0.003 | -0.007 | 0.144 | -0.016 | 0.002 |
| Parieto-occipital pontine (POPT) | -0.009 | 0.090 | -0.020 | 0.002 | -0.006 | 0.248 | -0.017 | 0.004 |
| Inferior cerebellar peduncle (ICP) | 0.005 | 0.484 | -0.009 | 0.018 | 0.003 | 0.561 | -0.008 | 0.015 |
| Superior cerebellar peduncle (SCP) | 0.005 | 0.437 | -0.007 | 0.017 | 0.004 | 0.467 | -0.007 | 0.015 |
| Inferior fronto occipital (IFO) | -0.008 | 0.171 | -0.019 | 0.003 | -0.008 | 0.157 | -0.019 | 0.003 |
| ST_ Fronto orbital (STFO) | -0.006 | 0.297 | -0.016 | 0.005 | -0.009 | 0.063 | -0.019 | 0.001 |
| ST_Occipital (STOCC) | -0.010 | 0.140 | -0.023 | 0.003 | -0.004 | 0.503 | -0.017 | 0.008 |
| ST_Parietal (STPAR) | -0.011 | 0.056 | -0.022 | 0.000 | -0.008 | 0.155 | -0.020 | 0.003 |
| ST_Postcentral (STPOSTC) | -0.011 | 0.061 | -0.022 | 0.001 | -0.007 | 0.251 | -0.019 | 0.005 |
| ST_Precentral (STPREC) | -0.011 | 0.055 | -0.022 | 0.000 | -0.009 | 0.138 | -0.020 | 0.003 |
| ST_Prefrontal (STPREF) | -0.007 | 0.130 | -0.017 | 0.002 | -0.009 | 0.069 | -0.018 | 0.001 |
| ST_Premotor (STPREM) | -0.007 | 0.152 | -0.017 | 0.003 | -0.008 | 0.116 | -0.018 | 0.002 |
| T_Occipital (TOCC) | -0.011 | 0.070 | -0.024 | 0.001 | -0.005 | 0.461 | -0.017 | 0.008 |
| T_Postcentral (TPOSTC) | -0.011 | 0.061 | -0.023 | 0.001 | -0.006 | 0.378 | -0.018 | 0.007 |
| T_Precentral (TPREC) | -.012* | 0.050 | -0.023 | 0.000 | -0.009 | 0.131 | -0.021 | 0.003 |
| T_Prefrontal (TPREF) | -0.008 | 0.112 | -0.017 | 0.002 | -0.008 | 0.078 | -0.018 | 0.001 |
| T_Premotor (TPREM) | -0.006 | 0.181 | -0.016 | 0.003 | -0.008 | 0.123 | -0.018 | 0.002 |
| Uncinate fascicle (UF) | -0.010 | 0.063 | -0.021 | 0.001 | -.014* | 0.009 | -0.024 | -0.004 |

**TW-RD**

| Tract | Left: Mean Difference | Left: *p* | Left: 95% CI - Lower | Left: 95% CI - Upper | Right: Mean Difference | Right: *p* | Right: 95% CI - Lower | Right: 95% CI - Upper |
|---|---|---|---|---|---|---|---|---|
| Anterior thalamic radiation (ATR) | 1.607E-5* | 0.018 | 2.939E-06 | 2.921E-05 | 2.066E-5* | 0.003 | 7.222E-06 | 3.411E-05 |
| Superior thalamic radiation (STR) | 2.142E-5* | 0.005 | 6.919E-06 | 3.592E-05 | 1.788E-5* | 0.018 | 3.236E-06 | 3.253E-05 |
| Corticospinal (CST) | 1.266E-5* | 0.046 | 2.258E-07 | 2.510E-05 | 1.211E-05 | 0.051 | -3.252E-08 | 2.426E-05 |
| Fronto pontine (FPT) | 1.324E-5* | 0.039 | 6.835E-07 | 2.579E-05 | 1.513E-5* | 0.019 | 2.624E-06 | 2.765E-05 |
| Parieto-occipital pontine (POPT) | 1.405E-5* | 0.031 | 1.387E-06 | 2.672E-05 | 1.070E-05 | 0.105 | -2.341E-06 | 2.375E-05 |
| Inferior cerebellar peduncle (ICP) | 1.431E-06 | 0.828 | -1.182E-05 | 1.468E-05 | 5.549E-06 | 0.413 | -7.996E-06 | 1.909E-05 |
| Superior cerebellar peduncle (SCP) | 7.733E-06 | 0.151 | -2.944E-06 | 1.841E-05 | 8.062E-06 | 0.151 | -3.071E-06 | 1.920E-05 |
| Inferior fronto occipital (IFO) | 1.595E-5* | 0.040 | 7.623E-07 | 3.114E-05 | 1.708E-5* | 0.031 | 1.618E-06 | 3.253E-05 |
| ST_ Fronto orbital (STFO) | 1.324E-05 | 0.072 | -1.264E-06 | 2.775E-05 | 1.768E-5* | 0.024 | 2.397E-06 | 3.295E-05 |
| ST_Occipital (STOCC) | 1.769E-05 | 0.050 | -3.867E-08 | 3.542E-05 | 1.731E-05 | 0.053 | -2.213E-07 | 3.485E-05 |
| ST_Parietal (STPAR) | 1.604E-5* | 0.029 | 1.737E-06 | 3.035E-05 | 1.346E-05 | 0.080 | -1.667E-06 | 2.858E-05 |
| ST_Postcentral (STPOSTC) | 1.753E-5* | 0.008 | 4.798E-06 | 3.026E-05 | 1.217E-05 | 0.096 | -2.237E-06 | 2.657E-05 |
| ST_Precentral (STPREC) | 1.906E-5* | 0.008 | 5.312E-06 | 3.281E-05 | 1.589E-5* | 0.031 | 1.493E-06 | 3.028E-05 |
| ST_Prefrontal (STPREF) | 1.553E-5* | 0.027 | 1.902E-06 | 2.917E-05 | 1.842E-5* | 0.012 | 4.364E-06 | 3.248E-05 |
| ST_Premotor (STPREM) | 1.578E-5* | 0.028 | 1.793E-06 | 2.977E-05 | 1.717E-5* | 0.018 | 3.098E-06 | 3.124E-05 |
| T_Occipital (TOCC) | 2.077E-5* | 0.030 | 2.137E-06 | 3.941E-05 | 2.017E-5* | 0.031 | 1.974E-06 | 3.837E-05 |
| T_Postcentral (TPOSTC) | 1.770E-5* | 0.006 | 5.393E-06 | 3.001E-05 | 1.184E-05 | 0.095 | -2.139E-06 | 2.582E-05 |
| T_Precentral (TPREC) | 1.971E-5* | 0.006 | 6.112E-06 | 3.331E-05 | 1.752E-5* | 0.016 | 3.410E-06 | 3.163E-05 |
| T_Prefrontal (TPREF) | 1.723E-5* | 0.014 | 3.725E-06 | 3.073E-05 | 1.967E-5* | 0.007 | 5.825E-06 | 3.352E-05 |
| T_Premotor (TPREM) | 1.651E-5* | 0.016 | 3.246E-06 | 2.978E-05 | 1.920E-5* | 0.006 | 5.856E-06 | 3.254E-05 |
| Uncinate fascicle (UF) | 1.638E-5* | 0.021 | 2.611E-06 | 3.015E-05 | 1.819E-5* | 0.020 | 3.064E-06 | 3.331E-05 |

**TW-AD**

| Tract | Left: Mean Difference | Left: *p* | Left: 95% CI - Lower | Left: 95% CI - Upper | Right: Mean Difference | Right: *p* | Right: 95% CI - Lower | Right: 95% CI - Upper |
|---|---|---|---|---|---|---|---|---|
| Anterior thalamic radiation (ATR) | 1.442E-05 | 0.102 | -2.970E-06 | 3.182E-05 | 1.364E-05 | 0.117 | -3.560E-06 | 3.083E-05 |
| Superior thalamic radiation (STR) | 1.199E-05 | 0.264 | -9.405E-06 | 3.339E-05 | 1.368E-05 | 0.191 | -7.095E-06 | 3.445E-05 |
| Corticospinal (CST) | 8.954E-06 | 0.305 | -8.465E-06 | 2.637E-05 | 7.983E-06 | 0.322 | -8.094E-06 | 2.406E-05 |
| Fronto pontine (FPT) | 9.196E-06 | 0.260 | -7.086E-06 | 2.548E-05 | 1.082E-05 | 0.158 | -4.363E-06 | 2.600E-05 |
| Parieto-occipital pontine (POPT) | 5.559E-06 | 0.485 | -1.037E-05 | 2.149E-05 | 6.166E-06 | 0.457 | -1.041E-05 | 2.274E-05 |
| Inferior cerebellar peduncle (ICP) | 9.894E-06 | 0.182 | -4.819E-06 | 2.461E-05 | 1.384E-05 | 0.073 | -1.328E-06 | 2.901E-05 |
| Superior cerebellar peduncle (SCP) | 2.199E-5* | 0.035 | 1.602E-06 | 4.237E-05 | 2.081E-5* | 0.033 | 1.803E-06 | 3.981E-05 |
| Inferior fronto occipital (IFO) | 1.024E-05 | 0.235 | -6.926E-06 | 2.740E-05 | 1.187E-05 | 0.169 | -5.246E-06 | 2.899E-05 |
| ST_ Fronto orbital (STFO) | 1.121E-05 | 0.209 | -6.552E-06 | 2.898E-05 | 1.087E-05 | 0.241 | -7.582E-06 | 2.932E-05 |
| ST_Occipital (STOCC) | 8.476E-06 | 0.446 | -1.376E-05 | 3.072E-05 | 1.911E-05 | 0.054 | -3.806E-07 | 3.860E-05 |
| ST_Parietal (STPAR) | 6.083E-06 | 0.454 | -1.017E-05 | 2.234E-05 | 6.046E-06 | 0.466 | -1.056E-05 | 2.265E-05 |
| ST_Postcentral (STPOSTC) | 1.158E-05 | 0.175 | -5.381E-06 | 2.855E-05 | 9.026E-06 | 0.316 | -8.938E-06 | 2.699E-05 |
| ST_Precentral (STPREC) | 1.403E-05 | 0.108 | -3.200E-06 | 3.125E-05 | 1.174E-05 | 0.170 | -5.231E-06 | 2.871E-05 |
| ST_Prefrontal (STPREF) | 1.178E-05 | 0.148 | -4.370E-06 | 2.794E-05 | 1.255E-05 | 0.127 | -3.722E-06 | 2.883E-05 |
| ST_Premotor (STPREM) | 1.585E-05 | 0.078 | -1.850E-06 | 3.355E-05 | 1.289E-05 | 0.118 | -3.437E-06 | 2.922E-05 |
| T_Occipital (TOCC) | 9.362E-06 | 0.431 | -1.442E-05 | 3.315E-05 | 2.189E-5* | 0.032 | 1.946E-06 | 4.184E-05 |
| T_Postcentral (TPOSTC) | 1.114E-05 | 0.196 | -5.974E-06 | 2.825E-05 | 1.128E-05 | 0.226 | -7.237E-06 | 2.979E-05 |
| T_Precentral (TPREC) | 1.325E-05 | 0.135 | -4.290E-06 | 3.080E-05 | 1.367E-05 | 0.112 | -3.322E-06 | 3.067E-05 |
| T_Prefrontal (TPREF) | 1.321E-05 | 0.117 | -3.441E-06 | 2.985E-05 | 1.450E-05 | 0.080 | -1.789E-06 | 3.078E-05 |
| T_Premotor (TPREM) | 1.707E-05 | 0.065 | -1.093E-06 | 3.524E-05 | 1.582E-05 | 0.052 | -1.420E-07 | 3.177E-05 |
| Uncinate fascicle (UF) | 8.823E-06 | 0.340 | -9.655E-06 | 2.730E-05 | 4.735E-06 | 0.644 | -1.585E-05 | 2.532E-05 |

Key: Mean difference: the mean difference of the "All patients" group minus the control group, based on the estimated marginal means of the univariate ANCOVA. Positive mean difference values indicate the "all patients" group had a higher average TW-TM than the controls, negative values indicate the control group had a higher average TW-TM than "all patients". "p" represents the significance of the mean difference.* a significant difference was observed in the estimated
