## Supplementary Table 3 for "White matter alterations in focal to bilateral tonic-clonic seizures" (from another version of this preprint: version 1, 2021-10-26)

**Supplementary Table 3.** Pairwise comparisons of the whole-tract, track-weighted mean differences between the FBTCS-N, FBTCS-Y and control groups.

| | | | TW-ADC | | | | TW-FA | | | | TW-RD | | | | TW-AD | | | |
|---|---|---|---|---|---|---|---|---|---|---|---|---|---|---|---|---|---|---|
| **Tracts** | **Group 1** | **Group 2** | **Mean Difference** | ***p*** | **95% CI Lower Bound** | **95% CI Upper Bound** | **Mean Difference** | ***p*** | **95% CI Lower Bound** | **95% CI Upper Bound** | **Mean Difference** | ***p*** | **95% CI Lower Bound** | **95% CI Upper Bound** | **Mean Difference** | ***p*** | **95% CI Lower Bound** | **95% CI Upper Bound** |
| Anterior thalamic radiation (ATR) | FBTCS-N | FBTCS-Y | 1.775E-05 | 0.214 | -6.225E-06 | 4.173E-05 | -0.010 | 0.397 | -0.027 | 0.006 | 1.835E-05 | 0.184 | -5.481E-06 | 4.218E-05 | 1.656E-05 | 0.576 | -1.465E-05 | 4.776E-05 |
| | | HC | **2.913E-5**$^{*}$ | **0.011** | **5.478E-06** | **5.278E-05** | -0.016 | 0.068 | -0.032 | 0.001 | **3.150E-5**$^{*}$ | **0.005** | **7.990E-06** | **5.501E-05** | 2.439E-05 | 0.163 | -6.387E-06 | 5.517E-05 |
| | HC | FBTCS-Y | -1.138E-05 | 0.339 | -2.893E-05 | 6.180E-06 | 0.005 | 0.838 | -0.007 | 0.017 | -1.315E-05 | 0.200 | -3.060E-05 | 4.304E-06 | -7.838E-06 | 1.000 | -3.069E-05 | 1.501E-05 |
| Superior thalamic radiation (STR) | FBTCS-N | FBTCS-Y | 1.001E-05 | 0.913 | -1.407E-05 | 3.409E-05 | -0.011 | 0.761 | -0.036 | 0.013 | 1.170E-05 | 0.838 | -1.501E-05 | 3.841E-05 | 6.627E-06 | 1.000 | -3.099E-05 | 4.425E-05 |
| | | HC | **2.408E-5**$^{*}$ | **0.046** | **3.239E-07** | **4.783E-05** | -0.019 | 0.156 | -0.043 | 0.005 | **2.814E-5**$^{*}$ | **0.033** | **1.792E-06** | **5.449E-05** | 1.595E-05 | 0.865 | -2.115E-05 | 5.306E-05 |
| | HC | FBTCS-Y | -1.407E-05 | 0.159 | -3.170E-05 | 3.565E-06 | 0.008 | 0.811 | -0.010 | 0.026 | -1.644E-05 | 0.126 | -3.599E-05 | 3.120E-06 | -9.326E-06 | 1.000 | -3.687E-05 | 1.822E-05 |
| Corticospinal (CST) | FBTCS-N | FBTCS-Y | 1.355E-05 | 0.274 | -6.042E-06 | 3.314E-05 | -0.008 | 0.908 | -0.028 | 0.012 | 1.301E-05 | 0.447 | -9.112E-06 | 3.514E-05 | 1.463E-05 | 0.655 | -1.464E-05 | 4.389E-05 |
| | | HC | **2.033E-5**$^{*}$ | **0.036** | **1.007E-06** | **3.966E-05** | -0.014 | 0.232 | -0.034 | 0.005 | **2.203E-5**$^{*}$ | **0.047** | **2.044E-07** | **4.385E-05** | 1.695E-05 | 0.449 | -1.192E-05 | 4.581E-05 |
| | HC | FBTCS-Y | -6.782E-06 | 0.731 | -2.113E-05 | 7.563E-06 | 0.006 | 0.945 | -0.009 | 0.020 | -9.015E-06 | 0.514 | -2.521E-05 | 7.184E-06 | -2.320E-06 | 1.000 | -2.374E-05 | 1.911E-05 |
| Fronto pontine (FPT) | FBTCS-N | FBTCS-Y | 1.689E-05 | 0.166 | -4.496E-06 | 3.828E-05 | -0.010 | 0.360 | -0.027 | 0.006 | 1.727E-05 | 0.188 | -5.288E-06 | 3.983E-05 | 1.613E-05 | 0.483 | -1.213E-05 | 4.438E-05 |
| | | HC | **2.446E-5**$^{*}$ | **0.018** | **3.364E-06** | **4.555E-05** | -0.015 | 0.079 | -0.031 | 0.001 | **2.671E-5**$^{*}$ | **0.014** | **4.459E-06** | **4.896E-05** | 1.995E-05 | 0.243 | -7.924E-06 | 4.782E-05 |
| | HC | FBTCS-Y | -7.568E-06 | 0.701 | -2.323E-05 | 8.090E-06 | 0.005 | 1.000 | -0.007 | 0.016 | -9.440E-06 | 0.481 | -2.596E-05 | 7.076E-06 | -3.823E-06 | 1.000 | -2.451E-05 | 1.687E-05 |
| Parieto-occipital pontine (POPT) | FBTCS-N | FBTCS-Y | 1.757E-05 | 0.105 | -2.540E-06 | 3.769E-05 | -0.009 | 0.812 | -0.028 | 0.011 | 1.611E-05 | 0.247 | -6.502E-06 | 3.871E-05 | 2.051E-05 | 0.179 | -5.945E-06 | 4.696E-05 |
| | | HC | **2.263E-5**$^{*}$ | **0.021** | **2.788E-06** | **4.246E-05** | -0.015 | 0.160 | -0.034 | 0.004 | **2.461E-5**$^{*}$ | **0.026** | **2.310E-06** | **4.691E-05** | 1.866E-05 | 0.244 | -7.432E-06 | 4.475E-05 |
| | HC | FBTCS-Y | -5.053E-06 | 1.000 | -1.978E-05 | 9.672E-06 | 0.007 | 0.760 | -0.008 | 0.021 | -8.504E-06 | 0.618 | -2.506E-05 | 8.049E-06 | 1.847E-06 | 1.000 | -1.752E-05 | 2.122E-05 |
| Inferior cerebellar peduncle (ICP) | FBTCS-N | FBTCS-Y | 7.696E-06 | 1.000 | -1.220E-05 | 2.759E-05 | -0.017 | 0.183 | -0.039 | 0.005 | 1.349E-05 | 0.426 | -9.049E-06 | 3.603E-05 | -3.897E-06 | 1.000 | -3.085E-05 | 2.305E-05 |
| | | HC | 1.350E-05 | 0.279 | -6.123E-06 | 3.313E-05 | -0.010 | 0.847 | -0.031 | 0.012 | 1.518E-05 | 0.286 | -7.051E-06 | 3.742E-05 | 1.015E-05 | 1.000 | -1.643E-05 | 3.673E-05 |
| | HC | FBTCS-Y | -5.809E-06 | 0.973 | -2.038E-05 | 8.760E-06 | -0.008 | 0.750 | -0.024 | 0.009 | -1.692E-06 | 1.000 | -1.820E-05 | 1.481E-05 | -1.405E-05 | 0.248 | -3.378E-05 | 5.687E-06 |
| Middle cerebellar peduncle (MCP) | FBTCS-N | FBTCS-Y | -8.252E-07 | 1.000 | -2.447E-05 | 2.282E-05 | -0.021 | 0.148 | -0.046 | 0.005 | 8.363E-06 | 1.000 | -1.821E-05 | 3.494E-05 | -1.920E-05 | 0.480 | -5.276E-05 | 1.436E-05 |
| | | HC | 7.628E-06 | 1.000 | -1.569E-05 | 3.095E-05 | -0.006 | 1.000 | -0.031 | 0.020 | 8.389E-06 | 1.000 | -1.782E-05 | 3.460E-05 | 6.106E-06 | 1.000 | -2.699E-05 | 3.921E-05 |
| | HC | FBTCS-Y | -8.454E-06 | 0.686 | -2.576E-05 | 8.858E-06 | -0.015 | 0.154 | -0.034 | 0.004 | -2.659E-08 | 1.000 | -1.948E-05 | 1.943E-05 | **-2.531E-5**$^{*}$ | **0.042** | **-4.988E-05** | **-7.365E-07** |
| Superior cerebellar peduncle (SCP) | FBTCS-N | FBTCS-Y | 3.882E-06 | 1.000 | -1.655E-05 | 2.431E-05 | -0.019 | 0.072 | -0.039 | 0.001 | 1.157E-05 | 0.410 | -7.490E-06 | 3.064E-05 | -1.150E-05 | 1.000 | -4.800E-05 | 2.500E-05 |
| | | HC | 1.569E-05 | 0.176 | -4.458E-06 | 3.584E-05 | -0.010 | 0.600 | -0.030 | 0.009 | 1.735E-05 | 0.079 | -1.455E-06 | 3.615E-05 | 1.238E-05 | 1.000 | -2.362E-05 | 4.838E-05 |
| | HC | FBTCS-Y | -1.181E-05 | 0.166 | -2.677E-05 | 3.147E-06 | -0.008 | 0.460 | -0.023 | 0.006 | -5.775E-06 | 0.920 | -1.973E-05 | 8.183E-06 | -2.388E-05 | 0.093 | -5.061E-05 | 2.844E-06 |
| Inferior fronto occipital (IFO) | FBTCS-N | FBTCS-Y | **2.502E-5**$^{*}$ | **0.044** | **5.022E-07** | **4.953E-05** | -0.010 | 0.574 | -0.030 | 0.009 | 2.318E-05 | 0.102 | -3.230E-06 | 4.958E-05 | **2.870E-5**$^{*}$ | **0.049** | **9.479E-08** | **5.731E-05** |
| | | HC | **3.247E-5**$^{*}$ | **0.005** | **8.291E-06** | **5.665E-05** | -0.016 | 0.129 | -0.036 | 0.003 | **3.344E-5**$^{*}$ | **0.008** | **7.390E-06** | **5.948E-05** | **3.055E-5**$^{*}$ | **0.030** | **2.331E-06** | **5.877E-05** |
| | HC | FBTCS-Y | -7.455E-06 | 0.914 | -2.540E-05 | 1.049E-05 | 0.006 | 0.964 | -0.009 | 0.020 | -1.026E-05 | 0.575 | -2.959E-05 | 9.073E-06 | -1.846E-06 | 1.000 | -2.279E-05 | 1.910E-05 |
| ST_ Fronto orbital (STFO) | FBTCS-N | FBTCS-Y | 1.630E-05 | 0.379 | -9.818E-06 | 4.241E-05 | -0.009 | 0.682 | -0.028 | 0.010 | 1.623E-05 | 0.423 | -1.080E-05 | 4.325E-05 | 1.644E-05 | 0.637 | -1.603E-05 | 4.890E-05 |
| | | HC | 2.542E-05 | 0.054 | -3.393E-07 | 5.118E-05 | -0.014 | 0.191 | -0.033 | 0.004 | **2.690E-5**$^{*}$ | **0.047** | **2.381E-07** | **5.355E-05** | 2.247E-05 | 0.261 | -9.547E-06 | 5.449E-05 |
| | HC | FBTCS-Y | -9.123E-06 | 0.719 | -2.824E-05 | 9.999E-06 | 0.005 | 1.000 | -0.009 | 0.019 | -1.067E-05 | 0.555 | -3.045E-05 | 9.120E-06 | -6.035E-06 | 1.000 | -2.980E-05 | 1.773E-05 |
| ST_Occipital (STOCC) | FBTCS-N | FBTCS-Y | **2.865E-5**$^{*}$ | **0.032** | **1.995E-06** | **5.531E-05** | -0.012 | 0.524 | -0.035 | 0.010 | 2.652E-05 | 0.091 | -3.016E-06 | 5.605E-05 | **3.292E-5**$^{*}$ | **0.033** | **2.084E-06** | **6.376E-05** |
| | | HC | **3.628E-5**$^{*}$ | **0.004** | **9.981E-06** | **6.257E-05** | -0.017 | 0.177 | -0.039 | 0.005 | **3.674E-5**$^{*}$ | **0.009** | **7.610E-06** | **6.587E-05** | **3.534E-5**$^{*}$ | **0.018** | **4.925E-06** | **6.576E-05** |
| | HC | FBTCS-Y | -7.622E-06 | 1.000 | -2.714E-05 | 1.190E-05 | 0.005 | 1.000 | -0.012 | 0.021 | -1.022E-05 | 0.731 | -3.185E-05 | 1.140E-05 | -2.420E-06 | 1.000 | -2.500E-05 | 2.016E-05 |
| ST_Parietal (STPAR) | FBTCS-N | FBTCS-Y | 2.146E-05 | 0.077 | -1.703E-06 | 4.463E-05 | -0.009 | 0.826 | -0.030 | 0.012 | 1.962E-05 | 0.194 | -6.207E-06 | 4.545E-05 | 2.514E-05 | 0.081 | -2.259E-06 | 5.254E-05 |
| | | HC | **2.721E-5**$^{*}$ | **0.015** | **4.364E-06** | **5.007E-05** | -0.017 | 0.125 | -0.038 | 0.003 | **2.941E-5**$^{*}$ | **0.019** | **3.933E-06** | **5.489E-05** | 2.282E-05 | 0.123 | -4.207E-06 | 4.984E-05 |
| | HC | FBTCS-Y | -5.752E-06 | 1.000 | -2.271E-05 | 1.121E-05 | 0.008 | 0.578 | -0.007 | 0.023 | -9.787E-06 | 0.608 | -2.870E-05 | 9.124E-06 | 2.322E-06 | 1.000 | -1.774E-05 | 2.238E-05 |
| ST_Postcentral (STPOSTC)** | FBTCS-N | FBTCS-Y | 1.122E-05 | 0.644 | -1.105E-05 | 3.348E-05 | -0.007 | 1.000 | -0.028 | 0.015 | 1.006E-05 | 0.932 | -1.446E-05 | 3.458E-05 | 1.353E-05 | 0.846 | -1.752E-05 | 4.458E-05 |
| | | HC | 2.154E-05 | 0.056 | -4.222E-07 | 4.350E-05 | -0.015 | 0.293 | -0.036 | 0.007 | 2.271E-05 | 0.072 | -1.470E-06 | 4.690E-05 | 1.919E-05 | 0.374 | -1.143E-05 | 4.982E-05 |
| | HC | FBTCS-Y | -1.032E-05 | 0.363 | -2.663E-05 | 5.978E-06 | 0.008 | 0.650 | -0.008 | 0.024 | -1.266E-05 | 0.256 | -3.061E-05 | 5.295E-06 | -5.662E-06 | 1.000 | -2.839E-05 | 1.707E-05 |
| ST_Precentral (STPREC) | FBTCS-N | FBTCS-Y | 1.400E-05 | 0.422 | -9.298E-06 | 3.729E-05 | -0.009 | 0.887 | -0.029 | 0.012 | 1.368E-05 | 0.548 | -1.156E-05 | 3.892E-05 | 1.463E-05 | 0.704 | -1.572E-05 | 4.498E-05 |
| | | HC | **2.571E-5**$^{*}$ | **0.024** | **2.731E-06** | **4.869E-05** | -0.017 | 0.132 | -0.037 | 0.003 | **2.753E-5**$^{*}$ | **0.026** | **2.635E-06** | **5.243E-05** | 2.206E-05 | 0.218 | -7.881E-06 | 5.199E-05 |
| | HC | FBTCS-Y | -1.171E-05 | 0.281 | -2.877E-05 | 5.344E-06 | 0.008 | 0.548 | -0.007 | 0.023 | -1.385E-05 | 0.204 | -3.234E-05 | 4.626E-06 | -7.422E-06 | 1.000 | -2.964E-05 | 1.480E-05 |
| ST_Prefrontal (STPREF) | FBTCS-N | FBTCS-Y | 1.795E-05 | 0.219 | -6.418E-06 | 4.231E-05 | -0.010 | 0.493 | -0.027 | 0.007 | 1.792E-05 | 0.245 | -7.188E-06 | 4.304E-05 | 1.799E-05 | 0.394 | -1.124E-05 | 4.723E-05 |
| | | HC | **2.785E-5**$^{*}$ | **0.018** | **3.819E-06** | **5.189E-05** | -0.016 | 0.075 | -0.032 | 0.001 | **2.987E-5**$^{*}$ | **0.013** | **5.099E-06** | **5.464E-05** | 2.381E-05 | 0.136 | -5.021E-06 | 5.265E-05 |
| | HC | FBTCS-Y | -9.905E-06 | 0.517 | -2.774E-05 | 7.934E-06 | 0.006 | 0.705 | -0.006 | 0.018 | -1.195E-05 | 0.336 | -3.033E-05 | 6.441E-06 | -5.820E-06 | 1.000 | -2.722E-05 | 1.558E-05 |
| ST_Premotor (STPREM) | FBTCS-N | FBTCS-Y | 1.679E-05 | 0.294 | -7.986E-06 | 4.156E-05 | -0.011 | 0.374 | -0.028 | 0.006 | 1.762E-05 | 0.260 | -7.480E-06 | 4.272E-05 | 1.512E-05 | 0.670 | -1.548E-05 | 4.572E-05 |
| | | HC | **2.728E-5**$^{*}$ | **0.024** | **2.848E-06** | **5.172E-05** | -0.016 | 0.068 | -0.033 | 0.001 | **2.911E-5**$^{*}$ | **0.016** | **4.358E-06** | **5.387E-05** | 2.362E-05 | 0.172 | -6.560E-06 | 5.380E-05 |
| | HC | FBTCS-Y | -1.050E-05 | 0.466 | -2.863E-05 | 7.641E-06 | 0.005 | 0.895 | -0.007 | 0.018 | -1.150E-05 | 0.376 | -2.987E-05 | 6.880E-06 | -8.503E-06 | 1.000 | -3.091E-05 | 1.390E-05 |
| T_Occipital (TOCC) | FBTCS-N | FBTCS-Y | **2.842E-5**$^{*}$ | **0.042** | **7.677E-07** | **5.607E-05** | -0.013 | 0.400 | -0.034 | 0.008 | 2.715E-05 | 0.088 | -2.878E-06 | 5.718E-05 | 3.097E-05 | 0.061 | -1.027E-06 | 6.296E-05 |
| | | HC | **3.822E-5**$^{*}$ | **0.004** | **1.095E-05** | **6.550E-05** | -0.019 | 0.089 | -0.040 | 0.002 | **3.993E-5**$^{*}$ | **0.005** | **1.031E-05** | **6.955E-05** | **3.481E-5**$^{*}$ | **0.026** | **3.255E-06** | **6.637E-05** |
| | HC | FBTCS-Y | -9.803E-06 | 0.698 | -3.005E-05 | 1.044E-05 | 0.006 | 1.000 | -0.010 | 0.021 | -1.278E-05 | 0.461 | -3.477E-05 | 9.204E-06 | -3.846E-06 | 1.000 | -2.727E-05 | 1.958E-05 |
| T_Postcentral (TPOSTC) | FBTCS-N | FBTCS-Y | 1.158E-05 | 0.515 | -9.240E-06 | 3.241E-05 | -0.008 | 1.000 | -0.031 | 0.014 | 1.145E-05 | 0.691 | -1.208E-05 | 3.498E-05 | 1.185E-05 | 1.000 | -1.956E-05 | 4.326E-05 |
| | | HC | **2.198E-5**$^{*}$ | **0.032** | **1.445E-06** | **4.252E-05** | -0.016 | 0.263 | -0.038 | 0.007 | **2.376E-5**$^{*}$ | **0.043** | **5.505E-07** | **4.696E-05** | 1.844E-05 | 0.433 | -1.254E-05 | 4.943E-05 |
| | HC | FBTCS-Y | -1.040E-05 | 0.287 | -2.565E-05 | 4.845E-06 | 0.007 | 0.843 | -0.009 | 0.024 | -1.231E-05 | 0.245 | -2.953E-05 | 4.918E-06 | -6.594E-06 | 1.000 | -2.959E-05 | 1.640E-05 |
| T_Precentral (TPREC) | FBTCS-N | FBTCS-Y | 1.390E-05 | 0.396 | -8.714E-06 | 3.650E-05 | -0.010 | 0.727 | -0.031 | 0.011 | 1.448E-05 | 0.459 | -1.040E-05 | 3.937E-05 | 1.272E-05 | 0.913 | -1.787E-05 | 4.331E-05 |
| | | HC | **2.650E-5**$^{*}$ | **0.015** | **4.195E-06** | **4.880E-05** | -0.018 | 0.098 | -0.039 | 0.002 | **2.927E-5**$^{*}$ | **0.015** | **4.731E-06** | **5.381E-05** | 2.094E-05 | 0.271 | -9.231E-06 | 5.111E-05 |
| | HC | FBTCS-Y | -1.260E-05 | 0.193 | -2.915E-05 | 3.953E-06 | 0.008 | 0.541 | -0.007 | 0.024 | -1.479E-05 | 0.147 | -3.301E-05 | 3.429E-06 | -8.222E-06 | 1.000 | -3.062E-05 | 1.417E-05 |
| T_Prefrontal (TPREF) | FBTCS-N | FBTCS-Y | 1.839E-05 | 0.187 | -5.582E-06 | 4.235E-05 | -0.011 | 0.349 | -0.028 | 0.006 | 1.908E-05 | 0.177 | -5.476E-06 | 4.363E-05 | 1.699E-05 | 0.475 | -1.259E-05 | 4.656E-05 |
| | | HC | **2.958E-5**$^{*}$ | **0.010** | **5.941E-06** | **5.322E-05** | **-.017**$^{*}$ | **0.048** | **-0.033** | **0.000** | **3.217E-5**$^{*}$ | **0.006** | **7.951E-06** | **5.639E-05** | 2.440E-05 | 0.129 | -4.774E-06 | 5.357E-05 |
| | HC | FBTCS-Y | -1.120E-05 | 0.355 | -2.874E-05 | 6.352E-06 | 0.006 | 0.715 | -0.006 | 0.018 | -1.309E-05 | 0.228 | -3.107E-05 | 4.886E-06 | -7.411E-06 | 1.000 | -2.907E-05 | 1.424E-05 |
| T_Premotor (TPREM) | FBTCS-N | FBTCS-Y | 1.729E-05 | 0.212 | -5.994E-06 | 4.057E-05 | -0.014 | 0.157 | -0.030 | 0.003 | 1.981E-05 | 0.121 | -3.557E-06 | 4.318E-05 | 1.226E-05 | 0.969 | -1.840E-05 | 4.291E-05 |
| | | HC | **2.917E-5**$^{*}$ | **0.009** | **6.206E-06** | **5.214E-05** | **-.018**$^{*}$ | **0.031** | **-0.035** | **-0.001** | **3.215E-5**$^{*}$ | **0.004** | **9.106E-06** | **5.520E-05** | 2.322E-05 | 0.186 | -7.018E-06 | 5.345E-05 |
| | HC | FBTCS-Y | -1.188E-05 | 0.267 | -2.893E-05 | 5.165E-06 | 0.004 | 1.000 | -0.008 | 0.017 | -1.234E-05 | 0.236 | -2.945E-05 | 4.764E-06 | -1.096E-05 | 0.686 | -3.341E-05 | 1.148E-05 |
| Uncinate fascicle (UF) | FBTCS-N | FBTCS-Y | 1.687E-05 | 0.341 | -9.223E-06 | 4.296E-05 | -0.001 | 1.000 | -0.020 | 0.018 | 1.159E-05 | 0.826 | -1.463E-05 | 3.780E-05 | 2.739E-05 | 0.163 | -7.156E-06 | 6.193E-05 |
| | | HC | 2.542E-05 | 0.054 | -3.147E-07 | 5.116E-05 | -0.012 | 0.338 | -0.031 | 0.007 | 2.508E-05 | 0.060 | -7.819E-07 | 5.094E-05 | 2.607E-05 | 0.189 | -7.999E-06 | 6.014E-05 |
| | HC | FBTCS-Y | -8.553E-06 | 0.808 | -2.766E-05 | 1.055E-05 | 0.012 | 0.133 | -0.002 | 0.025 | -1.349E-05 | 0.259 | -3.269E-05 | 5.703E-06 | 1.314E-06 | 1.000 | -2.398E-05 | 2.660E-05 |
| Commissure anterior (CA) | FBTCS-N | FBTCS-Y | 1.305E-05 | 0.672 | -1.340E-05 | 3.951E-05 | -0.014 | 0.508 | -0.040 | 0.011 | 1.632E-05 | 0.553 | -1.390E-05 | 4.654E-05 | 6.524E-06 | 1.000 | -3.029E-05 | 4.334E-05 |
| | | HC | 1.884E-05 | 0.236 | -7.257E-06 | 4.494E-05 | -0.014 | 0.561 | -0.039 | 0.012 | 2.053E-05 | 0.278 | -9.275E-06 | 5.034E-05 | 1.545E-05 | 0.880 | -2.086E-05 | 5.176E-05 |
| | HC | FBTCS-Y | -5.786E-06 | 1.000 | -2.516E-05 | 1.359E-05 | -0.001 | 1.000 | -0.020 | 0.018 | -4.215E-06 | 1.000 | -2.634E-05 | 1.791E-05 | -8.926E-06 | 1.000 | -3.588E-05 | 1.803E-05 |
| Corpus callosum (CC) | FBTCS-N | FBTCS-Y | 2.202E-05 | 0.075 | -1.600E-06 | 4.563E-05 | -0.011 | 0.387 | -0.030 | 0.007 | 2.208E-05 | 0.110 | -3.463E-06 | 4.762E-05 | 2.189E-05 | 0.133 | -4.479E-06 | 4.827E-05 |
| | | HC | **2.852E-5**$^{*}$ | **0.012** | **5.221E-06** | **5.181E-05** | **-.019**$^{*}$ | **0.043** | **-0.037** | **0.000** | **3.203E-5**$^{*}$ | **0.009** | **6.837E-06** | **5.723E-05** | 2.148E-05 | 0.136 | -4.528E-06 | 4.750E-05 |
| | HC | FBTCS-Y | -6.499E-06 | 1.000 | -2.379E-05 | 1.079E-05 | 0.007 | 0.564 | -0.006 | 0.021 | -9.952E-06 | 0.572 | -2.865E-05 | 8.750E-06 | 4.087E-07 | 1.000 | -1.890E-05 | 1.972E-05 |

Key: HC: control group; Mean difference: the difference between group 1 and group 2, based on the estimated marginal means of the univariate ANCOVA (Bonferroni correction selected). Positive mean difference values indicate group 1 had a higher average TW-TM than group 2, negative values indicate group 2 had a higher average TW-TM than group 1. "*p*" represents the significance of the mean difference.*a significant difference was observed in the estimated marginal means univariate ANCOVA output; **on the TW-ADC metric, there was a significant difference observed in the estimated marginal means of the univariate ANCOVA ($p < 0.047$), however the pairwise comparisons were non-significant.
