## Supplementary Table 4 for "White matter alterations in focal to bilateral tonic-clonic seizures" (from another version of this preprint: version 1, 2021-10-26)

**Supplementary Table 4.** Pairwise comparisons of the track-weighted, mean differences between the FBTCS-N, FBTCS-Y and control groups for each left and right side of a given tract.

| | | | TW-ADC | | | | | | | |
|---|---|---|---|---|---|---|---|---|---|---|
| | | | LEFT | | | | RIGHT | | | |
| **Tract** | **Group 1** | **Group 2** | **Mean Difference** | ***p*** | **95% CI Lower** | **95% CI Upper** | **Mean Difference** | ***p*** | **95% CI Lower** | **95% CI Upper** |
| Anterior thalamic radiation (ATR) | FBTCS-N | FBTCS-Y | 1.696E-05 | 0.258 | -7.150E-06 | 4.107E-05 | 1.842E-05 | 0.203 | -6.094E-06 | 4.294E-05 |
| | | HC | **2.713E-5**[*] | **0.021** | **3.354E-06** | **5.091E-05** | **3.119E-5**[*] | **0.008** | **7.009E-06** | **5.537E-05** |
| | HC | FBTCS-Y | -1.017E-05 | 0.471 | -2.783E-05 | 7.476E-06 | -1.277E-05 | 0.248 | -3.072E-05 | 5.181E-06 |
| Superior thalamic radiation (STR) | FBTCS-N | FBTCS-Y | 1.201E-05 | 0.664 | -1.218E-05 | 3.619E-05 | 8.251E-06 | 1.000 | -1.726E-05 | 3.376E-05 |
| | | HC | **2.566E-5**[*] | **0.031** | **1.804E-06** | **4.951E-05** | 2.282E-05 | 0.087 | -2.347E-06 | 4.798E-05 |
| | HC | FBTCS-Y | -1.365E-05 | 0.183 | -3.136E-05 | 4.054E-06 | -1.457E-05 | 0.175 | -3.325E-05 | 4.113E-06 |
| Corticospinal (CST) | FBTCS-N | FBTCS-Y | 1.505E-05 | 0.197 | -4.841E-06 | 3.494E-05 | 1.177E-05 | 0.490 | -8.969E-06 | 3.251E-05 |
| | | HC | **2.138E-5**[*] | **0.029** | **1.758E-06** | **4.100E-05** | 1.919E-05 | 0.072 | -1.267E-06 | 3.965E-05 |
| | HC | FBTCS-Y | -6.328E-06 | 0.850 | -2.089E-05 | 8.235E-06 | -7.419E-06 | 0.686 | -2.260E-05 | 7.766E-06 |
| Fronto pontine (FPT) | FBTCS-N | FBTCS-Y | 1.654E-05 | 0.193 | -5.206E-06 | 3.830E-05 | 1.720E-05 | 0.156 | -4.288E-06 | 3.869E-05 |
| | | HC | **2.309E-5**[*] | **0.031** | **1.635E-06** | **4.454E-05** | **2.590E-5**[*] | **0.012** | **4.706E-06** | **4.710E-05** |
| | HC | FBTCS-Y | -6.545E-06 | 0.930 | -2.247E-05 | 9.380E-06 | -8.701E-06 | 0.522 | -2.443E-05 | 7.033E-06 |
| Parieto-occipital pontine (POPT) | FBTCS-N | FBTCS-Y | 1.724E-05 | 0.127 | -3.321E-06 | 3.779E-05 | 1.705E-05 | 0.191 | -5.319E-06 | 3.943E-05 |
| | | HC | **2.293E-5**[*] | **0.022** | **2.657E-06** | **4.321E-05** | **2.230E-5**[*] | **0.047** | **2.322E-07** | **4.437E-05** |
| | HC | FBTCS-Y | -5.697E-06 | 1.000 | -2.075E-05 | 9.353E-06 | -5.246E-06 | 1.000 | -2.163E-05 | 1.113E-05 |
| Inferior cerebellar peduncle (ICP) | FBTCS-N | FBTCS-Y | 5.931E-06 | 1.000 | -1.340E-05 | 2.526E-05 | 9.522E-06 | 0.893 | -1.306E-05 | 3.210E-05 |
| | | HC | 1.048E-05 | 0.530 | -8.584E-06 | 2.955E-05 | 1.678E-05 | 0.201 | -5.498E-06 | 3.905E-05 |
| | HC | FBTCS-Y | -4.551E-06 | 1.000 | -1.870E-05 | 9.601E-06 | -7.253E-06 | 0.836 | -2.379E-05 | 9.280E-06 |
| Superior cerebellar peduncle (SCP) | FBTCS-N | FBTCS-Y | 2.142E-06 | 1.000 | -1.836E-05 | 2.265E-05 | 5.616E-06 | 1.000 | -1.577E-05 | 2.700E-05 |
| | | HC | 1.453E-05 | 0.240 | -5.698E-06 | 3.475E-05 | 1.697E-05 | 0.153 | -4.116E-06 | 3.806E-05 |
| | HC | FBTCS-Y | -1.239E-05 | 0.137 | -2.740E-05 | 2.627E-06 | -1.136E-05 | 0.231 | -2.701E-05 | 4.297E-06 |
| Inferior fronto occipital (IFO) | FBTCS-N | FBTCS-Y | 2.376E-05 | 0.073 | -1.636E-06 | 4.915E-05 | **2.606E-5**[*] | **0.047** | **2.252E-07** | **5.190E-05** |
| | | HC | **3.054E-5**[*] | **0.012** | **5.497E-06** | **5.559E-05** | **3.490E-5**[*] | **0.004** | **9.410E-06** | **6.038E-05** |
| | HC | FBTCS-Y | -6.787E-06 | 1.000 | -2.538E-05 | 1.181E-05 | -8.833E-06 | 0.748 | -2.775E-05 | 1.008E-05 |
| ST_ Fronto orbital (STFO) | FBTCS-N | FBTCS-Y | 1.286E-05 | 0.682 | -1.338E-05 | 3.910E-05 | 2.059E-05 | 0.200 | -6.720E-06 | 4.789E-05 |
| | | HC | 2.167E-05 | 0.128 | -4.220E-06 | 4.755E-05 | **3.031E-5**[*] | **0.023** | **3.380E-06** | **5.725E-05** |
| | HC | FBTCS-Y | -8.805E-06 | 0.775 | -2.802E-05 | 1.041E-05 | -9.727E-06 | 0.692 | -2.972E-05 | 1.027E-05 |
| ST_Occipital (STOCC) | FBTCS-N | FBTCS-Y | 2.958E-05 | 0.055 | -4.475E-07 | 5.962E-05 | 2.816E-05 | 0.059 | -8.203E-07 | 5.713E-05 |
| | | HC | **3.509E-5**[*] | **0.016** | **5.470E-06** | **6.471E-05** | **3.962E-5**[*] | **0.004** | **1.104E-05** | **6.820E-05** |
| | HC | FBTCS-Y | -5.508E-06 | 1.000 | -2.750E-05 | 1.648E-05 | -1.147E-05 | 0.551 | -3.268E-05 | 9.748E-06 |
| ST_Parietal (STPAR) | FBTCS-N | FBTCS-Y | 2.019E-05 | 0.117 | -3.473E-06 | 4.385E-05 | 2.206E-05 | 0.099 | -2.910E-06 | 4.702E-05 |
| | | HC | **2.678E-5**[*] | **0.020** | **3.442E-06** | **5.012E-05** | **2.770E-5**[*] | **0.023** | **3.074E-06** | **5.233E-05** |
| | HC | FBTCS-Y | -6.592E-06 | 1.000 | -2.392E-05 | 1.073E-05 | -5.643E-06 | 1.000 | -2.392E-05 | 1.264E-05 |
| ST_Postcentral (STPOSTC) | FBTCS-N | FBTCS-Y | 1.100E-05 | 0.641 | -1.077E-05 | 3.276E-05 | 1.104E-05 | 0.815 | -1.375E-05 | 3.583E-05 |
| | | HC | **2.282E-5**[*] | **0.034** | **1.345E-06** | **4.429E-05** | 1.987E-05 | 0.146 | -4.576E-06 | 4.433E-05 |
| | HC | FBTCS-Y | -1.182E-05 | 0.213 | -2.776E-05 | 4.117E-06 | -8.837E-06 | 0.691 | -2.699E-05 | 9.313E-06 |
| ST_Precentral (STPREC) | FBTCS-N | FBTCS-Y | 1.424E-05 | 0.415 | -9.328E-06 | 3.781E-05 | 1.332E-05 | 0.563 | -1.155E-05 | 3.820E-05 |
| | | HC | **2.668E-5**[*] | **0.020** | **3.436E-06** | **4.993E-05** | 2.441E-05 | 0.052 | -1.292E-07 | 4.894E-05 |
| | HC | FBTCS-Y | -1.244E-05 | 0.237 | -2.970E-05 | 4.814E-06 | -1.108E-05 | 0.407 | -2.929E-05 | 7.130E-06 |
| ST_Prefrontal (STPREF) | FBTCS-N | FBTCS-Y | 1.657E-05 | 0.287 | -7.722E-06 | 4.085E-05 | 1.923E-05 | 0.183 | -5.706E-06 | 4.416E-05 |
| | | HC | **2.571E-5**[*] | **0.032** | **1.751E-06** | **4.966E-05** | **3.005E-5**[*] | **0.012** | **5.457E-06** | **5.465E-05** |
| | HC | FBTCS-Y | -9.142E-06 | 0.617 | -2.692E-05 | 8.641E-06 | -1.082E-05 | 0.437 | -2.908E-05 | 7.433E-06 |
| ST_Premotor (STPREM) | FBTCS-N | FBTCS-Y | 1.707E-05 | 0.298 | -8.243E-06 | 4.238E-05 | 1.608E-05 | 0.356 | -9.144E-06 | 4.130E-05 |
| | | HC | **2.708E-5**[*] | **0.030** | **2.118E-06** | **5.205E-05** | **2.734E-5**[*] | **0.027** | **2.466E-06** | **5.222E-05** |
| | HC | FBTCS-Y | -1.002E-05 | 0.552 | -2.855E-05 | 8.516E-06 | -1.127E-05 | 0.404 | -2.973E-05 | 7.201E-06 |
| T_Occipital (TOCC) | FBTCS-N | FBTCS-Y | 3.116E-05 | 0.066 | -1.527E-06 | 6.384E-05 | 2.652E-05 | 0.106 | -3.925E-06 | 5.697E-05 |
| | | HC | **3.818E-5**[*] | **0.016** | **5.947E-06** | **7.042E-05** | **4.145E-5**[*] | **0.004** | **1.142E-05** | **7.148E-05** |
| | HC | FBTCS-Y | -7.029E-06 | 1.000 | -3.096E-05 | 1.690E-05 | -1.493E-05 | 0.305 | -3.722E-05 | 7.359E-06 |
| T_Postcentral (TPOSTC) | FBTCS-N | FBTCS-Y | 1.201E-05 | 0.445 | -8.387E-06 | 3.241E-05 | 1.065E-05 | 0.794 | -1.292E-05 | 3.423E-05 |
| | | HC | **2.353E-5**[*] | **0.017** | **3.412E-06** | **4.365E-05** | 2.020E-05 | 0.108 | -3.052E-06 | 4.345E-05 |
| | HC | FBTCS-Y | -1.152E-05 | 0.182 | -2.646E-05 | 3.414E-06 | -9.547E-06 | 0.522 | -2.681E-05 | 7.712E-06 |
| T_Precentral (TPREC) | FBTCS-N | FBTCS-Y | 1.514E-05 | 0.312 | -7.617E-06 | 3.789E-05 | 1.223E-05 | 0.641 | -1.198E-05 | 3.643E-05 |
| | | HC | **2.749E-5**[*] | **0.012** | **5.047E-06** | **4.993E-05** | **2.532E-5**[*] | **0.034** | **1.448E-06** | **4.920E-05** |
| | HC | FBTCS-Y | -1.235E-05 | 0.213 | -2.901E-05 | 4.305E-06 | -1.310E-05 | 0.216 | -3.082E-05 | 4.626E-06 |
| T_Prefrontal (TPREF) | FBTCS-N | FBTCS-Y | 1.778E-05 | 0.214 | -6.209E-06 | 4.177E-05 | 1.883E-05 | 0.187 | -5.724E-06 | 4.338E-05 |
| | | HC | **2.797E-5**[*] | **0.016** | **4.304E-06** | **5.163E-05** | **3.123E-5**[*] | **0.008** | **7.013E-06** | **5.545E-05** |
| | HC | FBTCS-Y | -1.019E-05 | 0.464 | -2.775E-05 | 7.377E-06 | -1.240E-05 | 0.276 | -3.038E-05 | 5.575E-06 |
| T_Premotor (TPREM) | FBTCS-N | FBTCS-Y | 1.761E-05 | 0.230 | -6.626E-06 | 4.184E-05 | 1.659E-05 | 0.250 | -6.774E-06 | 3.995E-05 |
| | | HC | **2.833E-5**[*] | **0.015** | **4.424E-06** | **5.223E-05** | **3.008E-5**[*] | **0.007** | **7.035E-06** | **5.313E-05** |
| | HC | FBTCS-Y | -1.072E-05 | 0.415 | -2.846E-05 | 7.023E-06 | -1.349E-05 | 0.167 | -3.060E-05 | 3.616E-06 |
| Uncinate fascicle (UF) | FBTCS-N | FBTCS-Y | 1.475E-05 | 0.445 | -1.028E-05 | 3.979E-05 | 1.884E-05 | 0.316 | -9.592E-06 | 4.728E-05 |
| | | HC | 2.441E-05 | 0.054 | -2.865E-07 | 4.911E-05 | 2.755E-05 | 0.056 | -4.956E-07 | 5.560E-05 |
| | HC | FBTCS-Y | -9.656E-06 | 0.585 | -2.799E-05 | 8.676E-06 | -8.708E-06 | 0.904 | -2.953E-05 | 1.211E-05 |

| | | | TW-FA | | | | | | | |
|---|---|---|---|---|---|---|---|---|---|---|
| | | | LEFT | | | | RIGHT | | | |
| **Tract** | **Group 1** | **Group 2** | **Mean Difference** | ***p*** | **95% CI Lower** | **95% CI Upper** | **Mean Difference** | ***p*** | **95% CI Lower** | **95% CI Upper** |
| Anterior thalamic radiation (ATR)* | FBTCS-N | FBTCS-Y | -0.012 | 0.247 | -0.030 | 0.005 | -0.008 | 0.729 | -0.024 | 0.009 |
| | | HC | -0.015 | 0.098 | -0.032 | 0.002 | -0.016 | 0.054 | -0.032 | 0.000 |
| | HC | FBTCS-Y | 0.003 | 1.000 | -0.010 | 0.015 | 0.008 | 0.285 | -0.004 | 0.020 |
| Superior thalamic radiation (STR) | FBTCS-N | FBTCS-Y | -0.007 | 1.000 | -0.033 | 0.018 | -0.015 | 0.413 | -0.039 | 0.010 |
| | | HC | -0.019 | 0.215 | -0.044 | 0.007 | -0.020 | 0.138 | -0.044 | 0.004 |
| | HC | FBTCS-Y | 0.011 | 0.414 | -0.007 | 0.030 | 0.005 | 1.000 | -0.013 | 0.023 |
| Corticospinal (CST) | FBTCS-N | FBTCS-Y | -0.008 | 1.000 | -0.029 | 0.013 | -0.009 | 0.793 | -0.028 | 0.011 |
| | | HC | -0.014 | 0.295 | -0.034 | 0.007 | -0.014 | 0.203 | -0.034 | 0.005 |
| | HC | FBTCS-Y | 0.006 | 0.986 | -0.009 | 0.021 | 0.006 | 0.991 | -0.009 | 0.020 |
| Fronto pontine (FPT) | FBTCS-N | FBTCS-Y | -0.009 | 0.570 | -0.026 | 0.008 | -0.012 | 0.220 | -0.028 | 0.004 |
| | | HC | -0.014 | 0.146 | -0.030 | 0.003 | **-.016*** | **0.042** | **-0.032** | **0.000** |
| | HC | FBTCS-Y | 0.005 | 1.000 | -0.008 | 0.017 | 0.004 | 1.000 | -0.007 | 0.016 |
| Parieto-occipital pontine (POPT) | FBTCS-N | FBTCS-Y | -0.008 | 0.977 | -0.028 | 0.012 | -0.010 | 0.641 | -0.029 | 0.009 |
| | | HC | -0.016 | 0.141 | -0.036 | 0.004 | -0.014 | 0.202 | -0.033 | 0.005 |
| | HC | FBTCS-Y | 0.008 | 0.502 | -0.006 | 0.023 | 0.004 | 1.000 | -0.009 | 0.018 |
| Inferior cerebellar peduncle (ICP) | FBTCS-N | FBTCS-Y | -0.019 | 0.168 | -0.043 | 0.005 | -0.016 | 0.210 | -0.037 | 0.005 |
| | | HC | -0.010 | 0.854 | -0.034 | 0.013 | -0.009 | 0.819 | -0.030 | 0.012 |
| | HC | FBTCS-Y | -0.009 | 0.680 | -0.026 | 0.009 | -0.006 | 0.905 | -0.022 | 0.009 |
| Superior cerebellar peduncle (SCP) | FBTCS-N | FBTCS-Y | -0.019 | 0.108 | -0.040 | 0.003 | **-.019*** | **0.048** | **-0.038** | **0.000** |
| | | HC | -0.010 | 0.780 | -0.031 | 0.012 | -0.011 | 0.439 | -0.030 | 0.008 |
| | HC | FBTCS-Y | -0.009 | 0.483 | -0.025 | 0.007 | -0.008 | 0.466 | -0.022 | 0.006 |
| Inferior fronto occipital (IFO) | FBTCS-N | FBTCS-Y | -0.012 | 0.455 | -0.033 | 0.009 | -0.008 | 0.878 | -0.028 | 0.011 |
| | | HC | -0.017 | 0.122 | -0.037 | 0.003 | -0.015 | 0.181 | -0.035 | 0.004 |
| | HC | FBTCS-Y | 0.005 | 1.000 | -0.010 | 0.020 | 0.007 | 0.770 | -0.008 | 0.021 |
| ST_ Fronto orbital (STFO) | FBTCS-N | FBTCS-Y | -0.011 | 0.530 | -0.031 | 0.009 | -0.007 | 1.000 | -0.026 | 0.012 |
| | | HC | -0.014 | 0.263 | -0.033 | 0.006 | -0.015 | 0.147 | -0.033 | 0.003 |
| | HC | FBTCS-Y | 0.003 | 1.000 | -0.012 | 0.017 | 0.008 | 0.473 | -0.006 | 0.022 |
| ST_Occipital (STOCC) | FBTCS-N | FBTCS-Y | -0.010 | 0.908 | -0.034 | 0.014 | -0.013 | 0.450 | -0.036 | 0.009 |
| | | HC | -0.018 | 0.203 | -0.042 | 0.006 | -0.015 | 0.290 | -0.038 | 0.007 |
| | HC | FBTCS-Y | 0.008 | 0.827 | -0.010 | 0.026 | 0.002 | 1.000 | -0.015 | 0.019 |
| ST_Parietal (STPAR) | FBTCS-N | FBTCS-Y | -0.009 | 0.904 | -0.030 | 0.012 | -0.010 | 0.786 | -0.031 | 0.012 |
| | | HC | -0.018 | 0.099 | -0.039 | 0.002 | -0.017 | 0.173 | -0.038 | 0.005 |
| | HC | FBTCS-Y | 0.009 | 0.390 | -0.006 | 0.025 | 0.007 | 0.859 | -0.009 | 0.022 |
| ST_Postcentral (STPOSTC) | FBTCS-N | FBTCS-Y | -0.007 | 1.000 | -0.028 | 0.015 | -0.006 | 1.000 | -0.029 | 0.017 |
| | | HC | -0.017 | 0.168 | -0.038 | 0.005 | -0.012 | 0.534 | -0.035 | 0.010 |
| | HC | FBTCS-Y | 0.010 | 0.377 | -0.006 | 0.026 | 0.006 | 1.000 | -0.011 | 0.023 |
| ST_Precentral (STPREC) | FBTCS-N | FBTCS-Y | -0.008 | 0.961 | -0.029 | 0.012 | -0.009 | 0.886 | -0.030 | 0.012 |
| | | HC | -0.018 | 0.110 | -0.038 | 0.003 | -0.016 | 0.201 | -0.037 | 0.005 |
| | HC | FBTCS-Y | 0.009 | 0.395 | -0.006 | 0.024 | 0.007 | 0.844 | -0.009 | 0.022 |
| ST_Prefrontal (STPREF) | FBTCS-N | FBTCS-Y | -0.010 | 0.502 | -0.027 | 0.008 | -0.009 | 0.526 | -0.026 | 0.008 |
| | | HC | -0.015 | 0.107 | -0.032 | 0.002 | -0.016 | 0.058 | -0.033 | 0.000 |
| | HC | FBTCS-Y | 0.005 | 0.952 | -0.008 | 0.018 | 0.007 | 0.503 | -0.005 | 0.019 |
| ST_Premotor (STPREM) | FBTCS-N | FBTCS-Y | -0.011 | 0.348 | -0.029 | 0.006 | -0.010 | 0.490 | -0.028 | 0.008 |
| | | HC | -0.016 | 0.083 | -0.034 | 0.001 | -0.017 | 0.075 | -0.034 | 0.001 |
| | HC | FBTCS-Y | 0.005 | 1.000 | -0.008 | 0.018 | 0.006 | 0.712 | -0.007 | 0.020 |
| T_Occipital (TOCC) | FBTCS-N | FBTCS-Y | -0.011 | 0.768 | -0.033 | 0.012 | -0.015 | 0.286 | -0.037 | 0.007 |
| | | HC | -0.020 | 0.096 | -0.043 | 0.002 | -0.017 | 0.165 | -0.039 | 0.005 |
| | HC | FBTCS-Y | 0.010 | 0.487 | -0.007 | 0.026 | 0.002 | 1.000 | -0.014 | 0.018 |
| T_Postcentral (TPOSTC) | FBTCS-N | FBTCS-Y | -0.008 | 1.000 | -0.030 | 0.014 | -0.009 | 1.000 | -0.032 | 0.015 |
| | | HC | -0.018 | 0.130 | -0.040 | 0.004 | -0.013 | 0.537 | -0.036 | 0.011 |
| | HC | FBTCS-Y | 0.010 | 0.395 | -0.006 | 0.026 | 0.004 | 1.000 | -0.013 | 0.021 |
| T_Precentral (TPREC) | FBTCS-N | FBTCS-Y | -0.009 | 0.907 | -0.030 | 0.012 | -0.011 | 0.609 | -0.033 | 0.010 |
| | | HC | -0.019 | 0.093 | -0.040 | 0.002 | -0.018 | 0.127 | -0.039 | 0.003 |
| | HC | FBTCS-Y | 0.010 | 0.361 | -0.006 | 0.026 | 0.007 | 0.890 | -0.009 | 0.022 |
| T_Prefrontal (TPREF) | FBTCS-N | FBTCS-Y | -0.011 | 0.372 | -0.028 | 0.006 | -0.011 | 0.365 | -0.027 | 0.006 |
| | | HC | -0.016 | 0.067 | -0.033 | 0.001 | **-.017*** | **0.040** | **-0.034** | **-0.001** |
| | HC | FBTCS-Y | 0.005 | 0.883 | -0.007 | 0.018 | 0.007 | 0.570 | -0.006 | 0.019 |
| T_Premotor (TPREM) | FBTCS-N | FBTCS-Y | -0.013 | 0.187 | -0.030 | 0.004 | -0.014 | 0.157 | -0.031 | 0.004 |
| | | HC | -0.017 | 0.052 | -0.034 | 0.000 | **-.019*** | **0.024** | **-0.036** | **-0.002** |
| | HC | FBTCS-Y | 0.004 | 1.000 | -0.009 | 0.016 | 0.005 | 0.915 | -0.007 | 0.018 |
| Uncinate fascicle (UF) | FBTCS-N | FBTCS-Y | -0.005 | 1.000 | -0.026 | 0.015 | 0.004 | 1.000 | -0.015 | 0.023 |
| | | HC | -0.014 | 0.267 | -0.034 | 0.006 | -0.011 | 0.462 | -0.030 | 0.008 |
| | HC | FBTCS-Y | 0.009 | 0.476 | -0.006 | 0.024 | **.015*** | **0.030** | **0.001** | **0.029** |

| | | | TW-RD | | | | | | | |
|---|---|---|---|---|---|---|---|---|---|---|
| | | | LEFT | | | | RIGHT | | | |
| Tract | Group 1 | Group 2 | Mean Difference | *p* | 95% CI Lower | 95% CI Upper | Mean Difference | *p* | 95% CI Lower | 95% CI Upper |
| Anterior thalamic radiation (ATR) | FBTCS-N | FBTCS-Y | 1.883E-05 | 0.165 | -4.989E-06 | 4.264E-05 | 1.763E-05 | 0.240 | -6.929E-06 | 4.218E-05 |
| | | HC | **2.946E-5*** | **0.010** | **5.967E-06** | **5.295E-05** | **3.363E-5*** | **0.004** | **9.414E-06** | **5.786E-05** |
| | HC | FBTCS-Y | -1.063E-05 | 0.405 | -2.807E-05 | 6.806E-06 | -1.601E-05 | 0.095 | -3.399E-05 | 1.971E-06 |
| Corticospinal (CST) | FBTCS-N | FBTCS-Y | 1.393E-05 | 0.413 | -9.074E-06 | 3.694E-05 | 1.199E-05 | 0.571 | -1.052E-05 | 3.451E-05 |
| | | HC | **2.272E-5*** | **0.050** | **2.566E-08** | **4.541E-05** | 2.129E-05 | 0.064 | -9.221E-07 | 4.349E-05 |
| | HC | FBTCS-Y | -8.786E-06 | 0.598 | -2.563E-05 | 8.057E-06 | -9.295E-06 | 0.498 | -2.578E-05 | 7.190E-06 |
| Fronto pontine (FPT) | FBTCS-N | FBTCS-Y | 1.612E-05 | 0.261 | -6.870E-06 | 3.911E-05 | 1.846E-05 | 0.142 | -4.100E-06 | 4.102E-05 |
| | | HC | **2.471E-5*** | **0.029** | **2.031E-06** | **4.739E-05** | **2.884E-5*** | **0.007** | **6.586E-06** | **5.109E-05** |
| | HC | FBTCS-Y | -8.587E-06 | 0.627 | -2.542E-05 | 8.247E-06 | -1.038E-05 | 0.371 | -2.689E-05 | 6.138E-06 |
| Inferior cerebellar peduncle (ICP) | FBTCS-N | FBTCS-Y | 1.278E-05 | 0.526 | -1.039E-05 | 3.594E-05 | 1.448E-05 | 0.410 | -9.363E-06 | 3.832E-05 |
| | | HC | 1.285E-05 | 0.501 | -9.999E-06 | 3.570E-05 | 1.802E-05 | 0.188 | -5.498E-06 | 4.154E-05 |
| | HC | FBTCS-Y | -7.573E-08 | 1.000 | -1.704E-05 | 1.689E-05 | -3.540E-06 | 1.000 | -2.100E-05 | 1.392E-05 |
| Inferior fronto occipital (IFO) | FBTCS-N | FBTCS-Y | 2.319E-05 | 0.121 | -4.159E-06 | 5.054E-05 | 2.280E-05 | 0.135 | -4.742E-06 | 5.034E-05 |
| | | HC | **3.251E-5*** | **0.014** | **5.534E-06** | **5.949E-05** | **3.470E-5*** | **0.008** | **7.537E-06** | **6.186E-05** |
| | HC | FBTCS-Y | -9.320E-06 | 0.753 | -2.934E-05 | 1.070E-05 | -1.190E-05 | 0.442 | -3.207E-05 | 8.259E-06 |
| Parieto-occipital pontine (POPT) | FBTCS-N | FBTCS-Y | 1.581E-05 | 0.290 | -7.432E-06 | 3.904E-05 | 1.596E-05 | 0.289 | -7.492E-06 | 3.941E-05 |
| | | HC | **2.565E-5*** | **0.024** | **2.726E-06** | **4.857E-05** | **2.349E-5*** | **0.045** | **3.589E-07** | **4.662E-05** |
| | HC | FBTCS-Y | -9.841E-06 | 0.467 | -2.685E-05 | 7.173E-06 | -7.531E-06 | 0.836 | -2.470E-05 | 9.637E-06 |
| Superior cerebellar peduncle (SCP) | FBTCS-N | FBTCS-Y | 9.832E-06 | 0.648 | -9.746E-06 | 2.941E-05 | 1.364E-05 | 0.276 | -6.125E-06 | 3.341E-05 |
| | | HC | 1.578E-05 | 0.143 | -3.526E-06 | 3.510E-05 | 1.925E-05 | 0.054 | -2.442E-07 | 3.875E-05 |
| | HC | FBTCS-Y | -5.953E-06 | 0.914 | -2.029E-05 | 8.381E-06 | -5.611E-06 | 1.000 | -2.008E-05 | 8.860E-06 |
| ST_ Fronto orbital (STFO) | FBTCS-N | FBTCS-Y | 1.421E-05 | 0.587 | -1.281E-05 | 4.123E-05 | 1.866E-05 | 0.313 | -9.416E-06 | 4.673E-05 |
| | | HC | 2.338E-05 | 0.102 | -3.269E-06 | 5.003E-05 | **3.140E-5*** | **0.022** | **3.706E-06** | **5.908E-05** |
| | HC | FBTCS-Y | -9.173E-06 | 0.758 | -2.896E-05 | 1.061E-05 | -1.274E-05 | 0.387 | -3.329E-05 | 7.815E-06 |
| ST_Occipital (STOCC) | FBTCS-N | FBTCS-Y | 2.617E-05 | 0.143 | -5.875E-06 | 5.821E-05 | 2.648E-05 | 0.114 | -4.387E-06 | 5.734E-05 |
| | | HC | **3.632E-5*** | **0.020** | **4.715E-06** | **6.793E-05** | **3.813E-5*** | **0.010** | **7.687E-06** | **6.858E-05** |
| | HC | FBTCS-Y | -1.015E-05 | 0.856 | -3.361E-05 | 1.331E-05 | -1.165E-05 | 0.613 | -3.425E-05 | 1.095E-05 |
| ST_Parietal (STPAR) | FBTCS-N | FBTCS-Y | 1.879E-05 | 0.240 | -7.358E-06 | 4.493E-05 | 2.000E-05 | 0.220 | -7.199E-06 | 4.719E-05 |
| | | HC | **2.978E-5*** | **0.019** | **3.993E-06** | **5.557E-05** | **2.910E-5*** | **0.030** | **2.279E-06** | **5.593E-05** |
| | HC | FBTCS-Y | -1.099E-05 | 0.475 | -3.014E-05 | 8.148E-06 | -9.107E-06 | 0.777 | -2.902E-05 | 1.081E-05 |
| ST_Postcentral (STPOSTC) | FBTCS-N | FBTCS-Y | 1.020E-05 | 0.876 | -1.370E-05 | 3.409E-05 | 9.636E-06 | 1.000 | -1.722E-05 | 3.649E-05 |
| | | HC | **2.489E-5*** | **0.035** | **1.322E-06** | **4.845E-05** | 2.014E-05 | 0.193 | -6.344E-06 | 4.663E-05 |
| | HC | FBTCS-Y | -1.469E-05 | 0.126 | -3.218E-05 | 2.800E-06 | -1.051E-05 | 0.566 | -3.017E-05 | 9.153E-06 |
| ST_Precentral (STPREC) | FBTCS-N | FBTCS-Y | 1.361E-05 | 0.572 | -1.197E-05 | 3.920E-05 | 1.344E-05 | 0.648 | -1.332E-05 | 4.020E-05 |
| | | HC | **2.858E-5*** | **0.022** | **3.350E-06** | **5.382E-05** | 2.617E-05 | 0.053 | -2.239E-07 | 5.257E-05 |
| | HC | FBTCS-Y | -1.497E-05 | 0.157 | -3.370E-05 | 3.760E-06 | -1.273E-05 | 0.336 | -3.232E-05 | 6.860E-06 |
| ST_Prefrontal (STPREF) | FBTCS-N | FBTCS-Y | 1.677E-05 | 0.306 | -8.289E-06 | 4.182E-05 | 1.895E-05 | 0.216 | -6.681E-06 | 4.458E-05 |
| | | HC | **2.750E-5*** | **0.025** | **2.785E-06** | **5.221E-05** | **3.230E-5*** | **0.008** | **7.019E-06** | **5.758E-05** |
| | HC | FBTCS-Y | -1.073E-05 | 0.453 | -2.908E-05 | 7.612E-06 | -1.335E-05 | 0.248 | -3.212E-05 | 5.416E-06 |
| ST_Premotor (STPREM) | FBTCS-N | FBTCS-Y | 1.786E-05 | 0.266 | -7.735E-06 | 4.346E-05 | 1.704E-05 | 0.318 | -8.735E-06 | 4.282E-05 |
| | | HC | **2.823E-5*** | **0.024** | **2.984E-06** | **5.348E-05** | **3.006E-5*** | **0.016** | **4.635E-06** | **5.549E-05** |
| | HC | FBTCS-Y | -1.037E-05 | 0.522 | -2.911E-05 | 8.370E-06 | -1.302E-05 | 0.277 | -3.189E-05 | 5.855E-06 |
| Superior thalamic radiation (STR) | FBTCS-N | FBTCS-Y | 1.063E-05 | 1.000 | -1.658E-05 | 3.784E-05 | 1.256E-05 | 0.767 | -1.469E-05 | 3.982E-05 |
| | | HC | **2.855E-5*** | **0.034** | **1.715E-06** | **5.539E-05** | **2.770E-5*** | **0.041** | **8.216E-07** | **5.459E-05** |
| | HC | FBTCS-Y | -1.792E-05 | 0.090 | -3.784E-05 | 2.001E-06 | -1.514E-05 | 0.195 | -3.510E-05 | 4.811E-06 |
| T_Occipital (TOCC) | FBTCS-N | FBTCS-Y | 2.793E-05 | 0.132 | -5.658E-06 | 6.152E-05 | 2.666E-05 | 0.129 | -5.251E-06 | 5.857E-05 |
| | | HC | **4.040E-5*** | **0.012** | **7.266E-06** | **7.353E-05** | **4.145E-5*** | **0.006** | **9.975E-06** | **7.293E-05** |
| | HC | FBTCS-Y | -1.247E-05 | 0.636 | -3.706E-05 | 1.213E-05 | -1.479E-05 | 0.363 | -3.816E-05 | 8.574E-06 |
| T_Postcentral (TPOSTC) | FBTCS-N | FBTCS-Y | 1.192E-05 | 0.602 | -1.101E-05 | 3.485E-05 | 1.068E-05 | 0.922 | -1.517E-05 | 3.653E-05 |
| | | HC | **2.643E-5*** | **0.017** | **3.806E-06** | **4.905E-05** | 2.075E-05 | 0.146 | -4.747E-06 | 4.624E-05 |
| | HC | FBTCS-Y | -1.451E-05 | 0.110 | -3.130E-05 | 2.284E-06 | -1.007E-05 | 0.572 | -2.899E-05 | 8.855E-06 |
| T_Precentral (TPREC) | FBTCS-N | FBTCS-Y | 1.486E-05 | 0.443 | -1.032E-05 | 4.004E-05 | 1.386E-05 | 0.576 | -1.228E-05 | 4.001E-05 |
| | | HC | **3.019E-5*** | **0.013** | **5.349E-06** | **5.503E-05** | **2.818E-5*** | **0.028** | **2.396E-06** | **5.397E-05** |
| | HC | FBTCS-Y | -1.533E-05 | 0.132 | -3.377E-05 | 3.108E-06 | -1.432E-05 | 0.206 | -3.346E-05 | 4.822E-06 |
| T_Prefrontal (TPREF) | FBTCS-N | FBTCS-Y | 1.849E-05 | 0.202 | -6.092E-06 | 4.308E-05 | 1.947E-05 | 0.179 | -5.651E-06 | 4.459E-05 |
| | | HC | **3.034E-5*** | **0.010** | **6.088E-06** | **5.459E-05** | **3.403E-5*** | **0.004** | **9.254E-06** | **5.881E-05** |
| | HC | FBTCS-Y | -1.185E-05 | 0.323 | -2.985E-05 | 6.156E-06 | -1.456E-05 | 0.164 | -3.296E-05 | 3.831E-06 |
| T_Premotor (TPREM) | FBTCS-N | FBTCS-Y | 1.962E-05 | 0.141 | -4.315E-06 | 4.356E-05 | 1.978E-05 | 0.135 | -4.115E-06 | 4.367E-05 |
| | | HC | **3.026E-5*** | **0.008** | **6.646E-06** | **5.387E-05** | **3.426E-5*** | **0.002** | **1.069E-05** | **5.782E-05** |
| | HC | FBTCS-Y | -1.063E-05 | 0.411 | -2.816E-05 | 6.893E-06 | -1.448E-05 | 0.135 | -3.198E-05 | 3.009E-06 |
| Uncinate fascicle (UF) | FBTCS-N | FBTCS-Y | 1.257E-05 | 0.685 | -1.315E-05 | 3.830E-05 | 1.055E-05 | 1.000 | -1.790E-05 | 3.901E-05 |
| | | HC | 2.527E-05 | 0.051 | -1.033E-07 | 5.064E-05 | 2.612E-05 | 0.075 | -1.946E-06 | 5.418E-05 |
| | HC | FBTCS-Y | -1.270E-05 | 0.299 | -3.153E-05 | 6.139E-06 | -1.557E-05 | 0.207 | -3.640E-05 | 5.266E-06 |

| | | | TW-AD | | | | | | | |
|---|---|---|---|---|---|---|---|---|---|---|
| | | | LEFT | | | | RIGHT | | | |
| Tract | Group 1 | Group 2 | Mean Difference | *p* | 95% CI Lower | 95% CI Upper | Mean Difference | *p* | 95% CI Lower | 95% CI Upper |
| Anterior thalamic radiation (ATR) | FBTCS-N | FBTCS-Y | 1.322E-05 | 0.932 | -1.901E-05 | 4.545E-05 | 2.001E-05 | 0.344 | -1.103E-05 | 5.106E-05 |
| | | HC | 2.248E-05 | 0.254 | -9.307E-06 | 5.427E-05 | 2.630E-05 | 0.114 | -4.319E-06 | 5.693E-05 |
| | HC | FBTCS-Y | -9.262E-06 | 0.995 | -3.286E-05 | 1.433E-05 | -6.292E-06 | 1.000 | -2.902E-05 | 1.644E-05 |
| Superior thalamic radiation (STR) | FBTCS-N | FBTCS-Y | 1.475E-05 | 1.000 | -2.419E-05 | 5.369E-05 | -3.687E-07 | 1.000 | -3.992E-05 | 3.918E-05 |
| | | HC | 1.987E-05 | 0.609 | -1.854E-05 | 5.828E-05 | 1.305E-05 | 1.000 | -2.596E-05 | 5.205E-05 |
| | HC | FBTCS-Y | -5.116E-06 | 1.000 | -3.363E-05 | 2.340E-05 | -1.341E-05 | 0.759 | -4.237E-05 | 1.554E-05 |
| Corticospinal (CST) | FBTCS-N | FBTCS-Y | 1.729E-05 | 0.502 | -1.348E-05 | 4.805E-05 | 1.133E-05 | 1.000 | -1.864E-05 | 4.130E-05 |
| | | HC | 1.870E-05 | 0.393 | -1.165E-05 | 4.904E-05 | 1.500E-05 | 0.634 | -1.456E-05 | 4.456E-05 |
| | HC | FBTCS-Y | -1.410E-06 | 1.000 | -2.394E-05 | 2.112E-05 | -3.668E-06 | 1.000 | -2.561E-05 | 1.827E-05 |
| Fronto pontine (FPT) | FBTCS-N | FBTCS-Y | 1.739E-05 | 0.439 | -1.197E-05 | 4.675E-05 | 1.469E-05 | 0.582 | -1.314E-05 | 4.252E-05 |
| | | HC | 1.985E-05 | 0.283 | -9.116E-06 | 4.881E-05 | 2.004E-05 | 0.226 | -7.413E-06 | 4.748E-05 |
| | HC | FBTCS-Y | -2.459E-06 | 1.000 | -2.396E-05 | 1.904E-05 | -5.347E-06 | 1.000 | -2.572E-05 | 1.503E-05 |
| Parieto-occipital pontine (POPT) | FBTCS-N | FBTCS-Y | 2.010E-05 | 0.229 | -7.526E-06 | 4.772E-05 | 1.924E-05 | 0.370 | -1.134E-05 | 4.983E-05 |
| | | HC | 1.751E-05 | 0.347 | -9.738E-06 | 4.475E-05 | 1.992E-05 | 0.319 | -1.025E-05 | 5.009E-05 |
| | HC | FBTCS-Y | 2.588E-06 | 1.000 | -1.764E-05 | 2.281E-05 | -6.728E-07 | 1.000 | -2.307E-05 | 2.172E-05 |
| Inferior cerebellar peduncle (ICP) | FBTCS-N | FBTCS-Y | -7.760E-06 | 1.000 | -3.507E-05 | 1.955E-05 | -3.925E-07 | 1.000 | -2.915E-05 | 2.836E-05 |
| | | HC | 5.743E-06 | 1.000 | -2.119E-05 | 3.268E-05 | 1.429E-05 | 0.644 | -1.408E-05 | 4.265E-05 |
| | HC | FBTCS-Y | -1.350E-05 | 0.297 | -3.350E-05 | 6.491E-06 | -1.468E-05 | 0.267 | -3.573E-05 | 6.375E-06 |
| Superior cerebellar peduncle (SCP) | FBTCS-N | FBTCS-Y | -1.324E-05 | 1.000 | -5.165E-05 | 2.518E-05 | -1.043E-05 | 1.000 | -4.622E-05 | 2.536E-05 |
| | | HC | 1.201E-05 | 1.000 | -2.588E-05 | 4.990E-05 | 1.242E-05 | 1.000 | -2.288E-05 | 4.772E-05 |
| | HC | FBTCS-Y | -2.525E-05 | 0.091 | -5.338E-05 | 2.874E-06 | -2.285E-05 | 0.106 | -4.905E-05 | 3.352E-06 |
| Inferior fronto occipital (IFO) | FBTCS-N | FBTCS-Y | 2.489E-05 | 0.145 | -5.660E-06 | 5.544E-05 | **3.259E-5**[*] | **0.028** | **2.794E-06** | **6.240E-05** |
| | | HC | 2.661E-05 | 0.099 | -3.523E-06 | 5.675E-05 | **3.529E-5**[*] | **0.014** | **5.894E-06** | **6.468E-05** |
| | HC | FBTCS-Y | -1.720E-06 | 1.000 | -2.409E-05 | 2.065E-05 | -2.694E-06 | 1.000 | -2.451E-05 | 1.913E-05 |
| ST_ Fronto orbital (STFO) | FBTCS-N | FBTCS-Y | 1.016E-05 | 1.000 | -2.340E-05 | 4.373E-05 | 2.445E-05 | 0.231 | -9.248E-06 | 5.814E-05 |
| | | HC | 1.824E-05 | 0.528 | -1.488E-05 | 5.135E-05 | 2.816E-05 | 0.121 | -5.080E-06 | 6.139E-05 |
| | HC | FBTCS-Y | -8.071E-06 | 1.000 | -3.265E-05 | 1.651E-05 | -3.708E-06 | 1.000 | -2.838E-05 | 2.096E-05 |
| ST_Occipital (STOCC) | FBTCS-N | FBTCS-Y | 3.642E-05 | 0.074 | -2.529E-06 | 7.537E-05 | 3.151E-05 | 0.086 | -3.189E-06 | 6.620E-05 |
| | | HC | 3.264E-05 | 0.120 | -5.779E-06 | 7.105E-05 | **4.260E-5**[*] | **0.010** | **8.379E-06** | **7.683E-05** |
| | HC | FBTCS-Y | 3.782E-06 | 1.000 | -2.473E-05 | 3.230E-05 | -1.110E-05 | 0.843 | -3.650E-05 | 1.431E-05 |
| ST_Parietal (STPAR) | FBTCS-N | FBTCS-Y | 2.300E-05 | 0.155 | -5.689E-06 | 5.168E-05 | 2.618E-05 | 0.102 | -3.606E-06 | 5.596E-05 |
| | | HC | 2.078E-05 | 0.221 | -7.511E-06 | 4.908E-05 | 2.489E-05 | 0.121 | -4.486E-06 | 5.427E-05 |
| | HC | FBTCS-Y | 2.213E-06 | 1.000 | -1.879E-05 | 2.322E-05 | 1.286E-06 | 1.000 | -2.052E-05 | 2.309E-05 |
| ST_Postcentral (STPOSTC) | FBTCS-N | FBTCS-Y | 1.260E-05 | 0.946 | -1.841E-05 | 4.361E-05 | 1.384E-05 | 0.931 | -1.988E-05 | 4.757E-05 |
| | | HC | 1.867E-05 | 0.404 | -1.192E-05 | 4.926E-05 | 1.933E-05 | 0.461 | -1.393E-05 | 5.260E-05 |
| | HC | FBTCS-Y | -6.076E-06 | 1.000 | -2.878E-05 | 1.663E-05 | -5.490E-06 | 1.000 | -3.018E-05 | 1.920E-05 |
| ST_Precentral (STPREC) | FBTCS-N | FBTCS-Y | 1.550E-05 | 0.654 | -1.550E-05 | 4.650E-05 | 1.309E-05 | 0.930 | -1.878E-05 | 4.497E-05 |
| | | HC | 2.288E-05 | 0.206 | -7.695E-06 | 5.346E-05 | 2.087E-05 | 0.314 | -1.056E-05 | 5.231E-05 |
| | HC | FBTCS-Y | -7.383E-06 | 1.000 | -3.008E-05 | 1.531E-05 | -7.781E-06 | 1.000 | -3.112E-05 | 1.555E-05 |
| ST_Prefrontal (STPREF) | FBTCS-N | FBTCS-Y | 1.616E-05 | 0.541 | -1.350E-05 | 4.583E-05 | 1.979E-05 | 0.306 | -9.793E-06 | 4.937E-05 |
| | | HC | 2.212E-05 | 0.198 | -7.136E-06 | 5.138E-05 | 2.555E-05 | 0.103 | -3.621E-06 | 5.473E-05 |
| | HC | FBTCS-Y | -5.957E-06 | 1.000 | -2.768E-05 | 1.576E-05 | -5.768E-06 | 1.000 | -2.743E-05 | 1.589E-05 |
| ST_Premotor (STPREM) | FBTCS-N | FBTCS-Y | 1.549E-05 | 0.701 | -1.656E-05 | 4.754E-05 | 1.415E-05 | 0.747 | -1.611E-05 | 4.440E-05 |
| | | HC | 2.479E-05 | 0.171 | -6.820E-06 | 5.641E-05 | 2.191E-05 | 0.221 | -7.933E-06 | 5.175E-05 |
| | HC | FBTCS-Y | -9.307E-06 | 0.980 | -3.277E-05 | 1.416E-05 | -7.762E-06 | 1.000 | -2.991E-05 | 1.439E-05 |
| T_Occipital (TOCC) | FBTCS-N | FBTCS-Y | 3.761E-05 | 0.087 | -3.851E-06 | 7.906E-05 | 2.624E-05 | 0.236 | -1.013E-05 | 6.261E-05 |
| | | HC | 3.376E-05 | 0.137 | -7.130E-06 | 7.465E-05 | **4.146E-5**[*] | **0.019** | **5.583E-06** | **7.733E-05** |
| | HC | FBTCS-Y | 3.844E-06 | 1.000 | -2.651E-05 | 3.420E-05 | -1.521E-05 | 0.482 | -4.184E-05 | 1.141E-05 |
| T_Postcentral (TPOSTC) | FBTCS-N | FBTCS-Y | 1.219E-05 | 0.999 | -1.895E-05 | 4.334E-05 | 1.060E-05 | 1.000 | -2.439E-05 | 4.559E-05 |
| | | HC | 1.774E-05 | 0.468 | -1.297E-05 | 4.846E-05 | 1.910E-05 | 0.521 | -1.541E-05 | 5.362E-05 |
| | HC | FBTCS-Y | -5.550E-06 | 1.000 | -2.835E-05 | 1.725E-05 | -8.503E-06 | 1.000 | -3.412E-05 | 1.712E-05 |
| T_Precentral (TPREC) | FBTCS-N | FBTCS-Y | 1.569E-05 | 0.657 | -1.576E-05 | 4.714E-05 | 8.954E-06 | 1.000 | -2.316E-05 | 4.107E-05 |
| | | HC | 2.209E-05 | 0.247 | -8.930E-06 | 5.311E-05 | 1.960E-05 | 0.388 | -1.207E-05 | 5.128E-05 |
| | HC | FBTCS-Y | -6.401E-06 | 1.000 | -2.943E-05 | 1.663E-05 | -1.065E-05 | 0.791 | -3.416E-05 | 1.286E-05 |
| T_Prefrontal (TPREF) | FBTCS-N | FBTCS-Y | 1.635E-05 | 0.553 | -1.394E-05 | 4.663E-05 | 1.754E-05 | 0.440 | -1.211E-05 | 4.720E-05 |
| | | HC | 2.322E-05 | 0.177 | -6.656E-06 | 5.309E-05 | 2.562E-05 | 0.103 | -3.626E-06 | 5.488E-05 |
| | HC | FBTCS-Y | -6.870E-06 | 1.000 | -2.904E-05 | 1.530E-05 | -8.080E-06 | 1.000 | -2.979E-05 | 1.363E-05 |
| T_Premotor (TPREM) | FBTCS-N | FBTCS-Y | 1.357E-05 | 0.926 | -1.937E-05 | 4.651E-05 | 1.022E-05 | 1.000 | -1.939E-05 | 3.982E-05 |
| | | HC | 2.446E-05 | 0.201 | -8.032E-06 | 5.696E-05 | 2.172E-05 | 0.211 | -7.480E-06 | 5.093E-05 |
| | HC | FBTCS-Y | -1.089E-05 | 0.796 | -3.501E-05 | 1.323E-05 | -1.150E-05 | 0.575 | -3.318E-05 | 1.017E-05 |
| Uncinate fascicle (UF) | FBTCS-N | FBTCS-Y | 1.911E-05 | 0.514 | -1.522E-05 | 5.344E-05 | 3.543E-05 | 0.059 | -1.026E-06 | 7.188E-05 |
| | | HC | 2.269E-05 | 0.304 | -1.117E-05 | 5.655E-05 | 3.042E-05 | 0.122 | -5.539E-06 | 6.638E-05 |
| | HC | FBTCS-Y | -3.578E-06 | 1.000 | -2.871E-05 | 2.155E-05 | 5.010E-06 | 1.000 | -2.168E-05 | 3.170E-05 |

Key: HC: control group; Mean difference: the difference between group 1 and group 2, based on the estimated marginal means of the univariate ANCOVA (Bonferroni correction selected). Positive mean difference values indicate group 1 had a higher average TW-TM than group 2, negative values indicate group 2 had a higher average TW-TM than group 1. "*p*" represents the significance of the mean difference.* a significant difference was observed in the estimated marginal means univariate ANCOVA. **for the right ATR tract, there was a significant difference observed in the estimated
